# Access to assistive products, home modifications and human help in older adults in the 2021-2025 Autonomy survey

**DOI:** 10.64898/2026.09.09.26362610

**Authors:** Gabriella C Silva, Aurore Fayosse, Louis Jacob, Séverine Sabia, Archana Singh-Manoux, Benjamin Landré

## Abstract

Limited evidence on access to assistive technologies among older adults may obscure significant inequities and hinder the development of effective policies aimed at reducing unmet need. This secondary analysis of the 2021-2025 Autonomy survey conducted in France used data from 8,024 respondents aged 60 and above on access to 54 assistive products across nine functional domains, 34 home modifications across 5 areas of the home and 24 activities requiring human help. Using survey weights, use, unmet need, total need, met need, undermet need and coverage were calculated across the total population and for vulnerable subgroups. Reasons for non-use were examined among those not using needed assistive products. We estimated that 83·2% (81·8%, 84·5%) of the French older population use assistive technology. Use was largest for assistive products for seeing, reading and for everyday life, home modifications in the bathing area and human help for undertaking less frequent tasks. Coverage was low for assistive products for communication and for transfers, home modifications in the kitchen and bathing area, as well as human help for running errands and using electronic devices. Use and unmet need were more than five times greater than the total population for subgroups with functional limitations. Cost was the biggest barrier to access. While the majority of total need for assistive technologies corresponded to use, important gaps in coverage were identified that merit attention to support an autonomous life among older populations.

## Introduction

Rapid population ageing is transforming societies and placing new demands on health and social care systems worldwide.^1,2^ Increases in life expectancy have been accompanied by a greater proportion of life lived with chronic diseases and functional limitations,^3^ shifting needs from acute medical care toward sustained support. As a result, the availability, coordination, and financing of technologies and services designed to support an autonomous, high-quality life among older adults must be prioritized.^4,5^ To accomplish this, public health policies have developed strategies that promote healthy ageing, functional ability, and ageing in place. Frameworks such as the World Health Organization’s Integrated Care for Older People emphasize the prevention of functional decline and the optimization of intrinsic capacity through integrated, person-centered approaches.^6^ A key component of these strategies is the availability and effective use of assistive technologies, comprised of assistive products, home modifications, and human help.^7^ These technologies can mitigate functional limitations, enhance autonomy, and support social participation among older adults.^8^

Current evidence suggests that access to assistive technologies remains uneven,^9–11^ and that access to specific categories of these resources are disproportionately impacted by individual and contextual barriers.^12–15^ Prior large scale studies rely mainly on analyses conducted in the general adult population,^10,16,17^ without a specific focus on older adults, whose needs and patterns of use may differ substantially. Furthermore, to our knowledge, there are no large-scale studies focusing on accessibility to assistive technologies among vulnerable subgroups of older adults, such as individuals with frailty, multimorbidity or limitations in activities of daily living, despite the likely existence of substantial heterogeneity in both needs and access. Failure to consider this heterogeneity may obscure critical inequities and lead to policy conclusions that are not applicable to the most vulnerable groups, thereby undermining effective resource allocation and intervention planning. In addition, much of the existing literature focuses on a narrow range of products and services and only reports utilization or need for these; doing so fails to capture the full extent to which these resources are used and other key dimensions of access such as coverage, or the extent to which identified needs are met by appropriate resources.^18^

Using data from the large, nationally representative 2021-2025 Autonomy survey conducted in France, this study aims to provide a comprehensive assessment of the use, need, and coverage of assistive products, home modifications and human assistance among older adults aged 60 years or more and across vulnerable population subgroups.

## Methods

### Study population

Data are drawn from the Autonomy survey^19^ undertaken in 2021-2025 by the French National Institute of Statistics and Economic Studies (Institut National de la Statistique et des Etudes Economiques, INSEE) and the French Directorate of Research, Studies, Assessment, and Statistics (Direction de la Recherche, des Etudes, de l’Evaluation et des Statistiques, DREES). Results from a preliminary survey, Vie Quotidienne et Santé, were used to select survey participants by oversampling people with disability. Participants were interviewed and INSEE and DRESS provided weights to adjust results from the Autonomy survey to be nationally representative. This secondary analysis of the survey data focuses on community-dwelling respondents 60 years and older.

### Access to assistive products, home modifications, and human help

Participants reported their use and need for assistive products, home modifications and human help. There were 54 assistive products across nine functional groups: 1. Seeing/reading; 2. Hearing/speaking; 3. Prosthetics and implants; 4. Orthoses; 5. Personal care; 6. Everyday life; 7. Personal mobility; 8. Transfers, getting up, and going to bed, and 9. Communication and to manage everyday life. Required home modifications were assessed using 34 items across 5 domains: 1. Toilet area, 2. Bathing area, 3. Kitchen, 4. Stairs, and 5. Other areas of the residence (**Table S1**). Human help consists of 24 activities potentially requiring human assistance (i.e. bathing, using the toilet, preparing meals, …) (**Table S2)**. Among participants specifying need for assistive products or home modifications that they did not have, the reasons for this were requested from the list shown in **Table S3** and included cost, difficulty to use, preference, etc. A comprehensive list of all assistive technologies considered is available is **Figure S1**.

### Covariates

Sociodemographic variables included age, sex, marital status, and education (low, intermediate, high). Participants reported whether they had foregone care in the past 12 months and, if so, the underlying reasons among cost, waiting time, lack of referral letter, etc. (details in **Table S4**). The presence of chronic conditions was assessed based on a predefined list of 47 chronic conditions (**Table S5**). Participants reporting several conditions were considered multimorbid. Frailty was defined as satisfying three or more criteria of Fried’s phenotype^20^ (slowness, weakness, exhaustion, low activity and weight loss), adapted to self-reported data.

We considered three measures of disability. <u>One</u>, we use Stineman’s classification of activities of daily living (ADL) and instrumental activities of daily living (IADL)^21^. ADL included eating, using the toilet, dressing, bathing, getting in and out of the bed/chair, and walking; IADL included using the telephone, money management, meal preparation, light housework, heavy housework, and shopping (**Table S6**). This framework defines five stages of limitations for both ADL and IADL but small numbers in stages I to IV led us to combine them in the analyses. <u>Two</u>, disability was assessed using the AGGIR (Autonomie Gérontologie Groupes Iso-Ressources) scale, an instrument used in France to assess a person’s autonomy and determine allowances granted by the government and the level of monitoring required for the person^22^. The six resulting GIR (Groupes Iso-Ressources) categories were grouped into two for analysis, with groups 1–4 combined as they qualify for additional government support. <u>Three</u>, a self-reported measure in which participants indicated whether a health problem had limited their activities during the previous six months.

### Statistical analysis

First, accessibility indicators were calculated using the definitions proposed by Danemayer et al.^17^. We defined six access indicators: 1. use (proportion of the population reporting use of a particular tool – an assistive product, home modification or human help), 2. total need (proportion of the population that either use or report needing the tool), 3. met need (proportion of the population whose need is fully satisfied by the tool they use), 4. undermet need (proportion of the population that use a tool but report needing an updated or different tool within the same group category), 5. unmet need (proportion of the population who reports needing a tool but does not use one), and 6. coverage (proportion of individuals among those using or needing a tool whose need is met) (**Supplementary Method 1**). Use, total need, and unmet need were derived at both the specific and group level for assistive products and home modifications. For assistive products and home modifications, met need, under-met need, and coverage were only calculated at the group level (with exception for glasses and hearing aids). All six access indicators could be calculated for the activities requiring human help. Weighted prevalence estimates and corresponding 95% confidence intervals (CIs) were computed for all accessibility indicators. Among those reporting needing an assistive product or home modification but not using any tool, we computed the prevalence for reasons for non-use.

Second, accessibility indicators were recalculated for subgroups of the population. The subgroups considered spanned three dimensions: sociodemographic (oldest-old (age ≥80 years), female, low education level, and participants renouncing use of medical care), morbidity (frail, participants with chronic conditions, and multimorbidity), limitations (ADL limitation (Stineman ADL Stage = I-IV), IADL limitation (Stineman IADL Stage = I-IV), any ADL or IADL limitation, a GIR categorization of 1 to 4, and self-reported limitations).

All analyses were conducted using R software (version 4.3.2). Survey analyses were conducted using the *survey* (version 4.4.8) and *srvyr* (version 1.3.0) packages, respectively. The research protocol was pre-registered; deviations from this protocol are mainly related to data availability (**Supplementary Method 2**).

## Results

### Study Population

We included 8,024 participants aged ≥60 years, representing 16,715,451 older adults living in France (**Table S7**). The weighted mean age of participants was 71·7 ± 0·13 years, with 19·4% (observed frequency, N=2,339) aged 80 years or older. The target population, the French population of older adults, were more likely to be female (55·1%, N=4,463) and to have a low education level (65·4%, N=5,826). Individuals who renounced care (9·4% (N=1,053)) during the last 12 months reported reasons related to cost (32·6%), other (27·9%) and long waiting times (13·5%) (**Table S8**). Frailty was present in 24·8% (N=4,049) of the total population and 79·9% (N=7,240) reported at least one chronic condition (**Table S7, S9**). Only 7·6% (N=1,679) of individuals were classified into GIR groups 1 through 4, 12·8% (N=2,498) reported a limitation in ADLs, 25·3% (N=3,817) reported IADL limitations and 28·0% (N=4,276) reported a limitation in either IADLs or ADLs. Most ADL and IADL limitations were considered Stage II (ADL: 7.1%, IADL: 12.7%); 38·1% (N=5,207) of the population reported having limitations in the past 6 months due to a health problem.

### Access Indicators – Total Population

For assistive products, 80·6% of the population used at least one product, 7·2% had undermet needs and 0·5% had unmet needs (**Table S10**). Among those who used or needed an assistive product, 90.5% reported using the appropriate tool. Home modifications were used by 21·9% of participants (undermet need 4·9%; unmet need 3·6%) (**Figure 1, Table S10**), coverage was 66·8% (**Figure S2**). Relying on human help to complete any of the 24 activities in **Table S2** was reported by 18·5% of the population (undermet need 5·1%; unmet need 3·1%) (**Figure 1, Table S10)**; coverage for human help was 62·2% (**Figure S2**). Across all assistive technologies (assistive products, home modifications, and human help), 83·2% used one of these tools, 18·5% had undermet need and 0·6% had unmet need, resulting in a coverage of 77·2%. Excluding glasses/contact lenses, used by over 75% of the population, considerably impacted these access indicators, decreasing use for assistive technologies to 46·5% and for assistive products to 35·7%.

**Figure 1.**
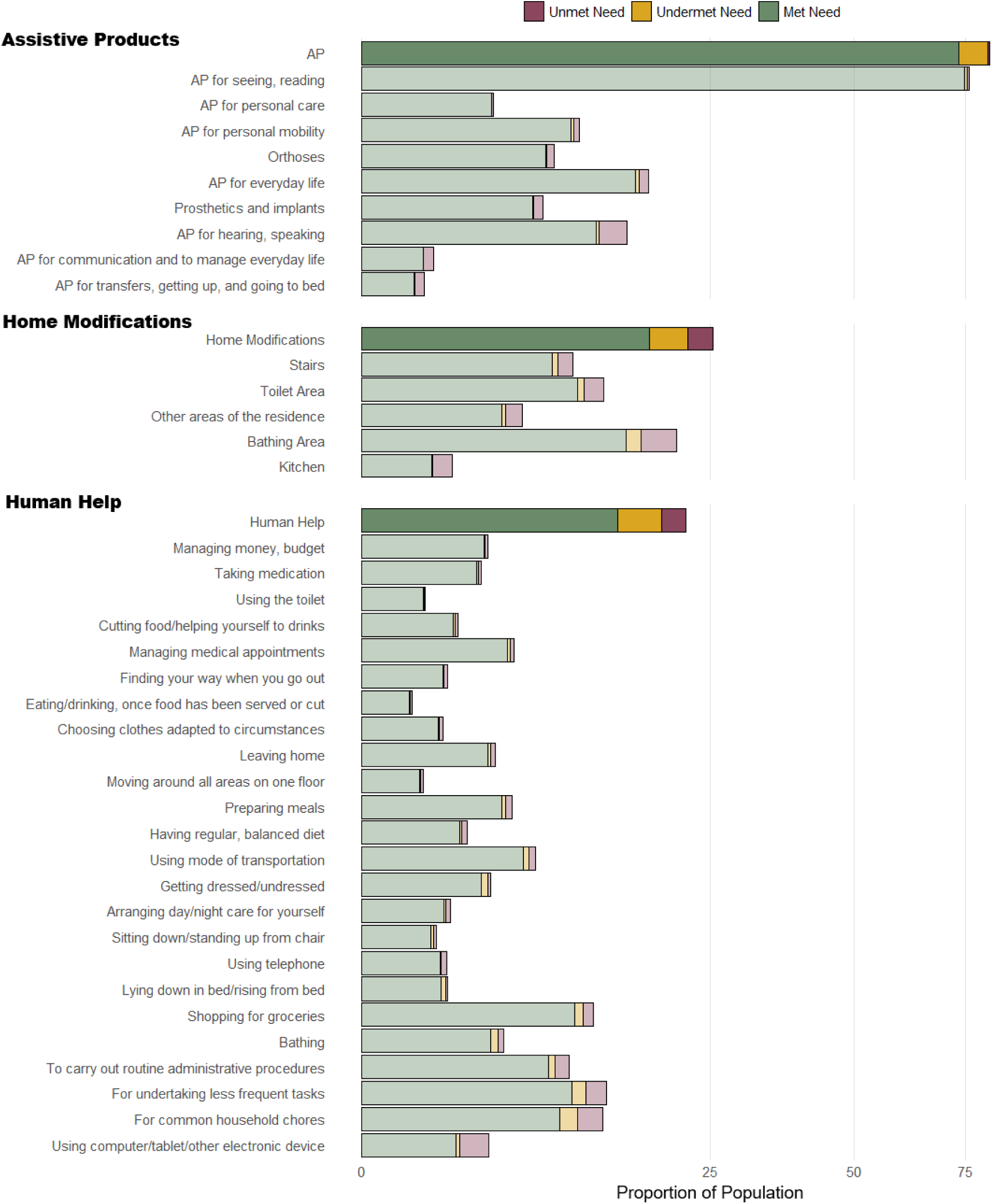
Proportion of Population with Unmet, Undermet and Met Need across Assistive Products, Home Modifications and Human Help. Abbreviations: AP: Assistive Products Met Need: the proportion of the population whose need is fully satisfied by the tool they use, Undermet Need: the proportion of the population that use a tool but report needing an updated or different tool within the same group category, Unmet Need: the proportion of the population who reports needing a tool but does not use one

When disaggregated to the level of assistive product category and device (**Figure 2**; **Table S10**), products for seeing and reading were by far the most frequently used (75·5%) and had the highest coverage (98·5%). This was largely driven by glasses/contact lenses, which accounted for 99·4% of devices in this category. Only 75·2% of those using or needing glasses/contacts reported having the appropriate device. The second and third assistive product categories most frequently used were tools for everyday life (use 15·9%; coverage 91·1%) and for hearing and speaking (use 11·6%; coverage 77·7%); within these categories, dentures and hearing aids represented 82·0% and 88·2% of devices used, respectively. Despite their high use, hearing aids and hearing implants had low coverage, 71·0% and 50·1%, respectively. Use and coverage were also relatively high for products for personal mobility (use 9·2%; coverage 92·7%), orthoses (7·0%; 91·4%), prosthetics and implants (6·1%; 88·9%), and products for personal care (3·5%; 96·9%). In contrast, products for transfers and for communication and managing everyday life were least frequently used (0·6% and 0·8%, respectively) and also showed comparatively lower coverage (70·1% and 72·9%, respectively). When participants were asked why they didn’t have assistive products they needed, cost (35.4%), preferences (15.6%), and challenges in obtaining one (9.8%) were the primary reasons (**Table S11**); 22.8% of these individuals reported “other” as the reason.

**Figure 2:**
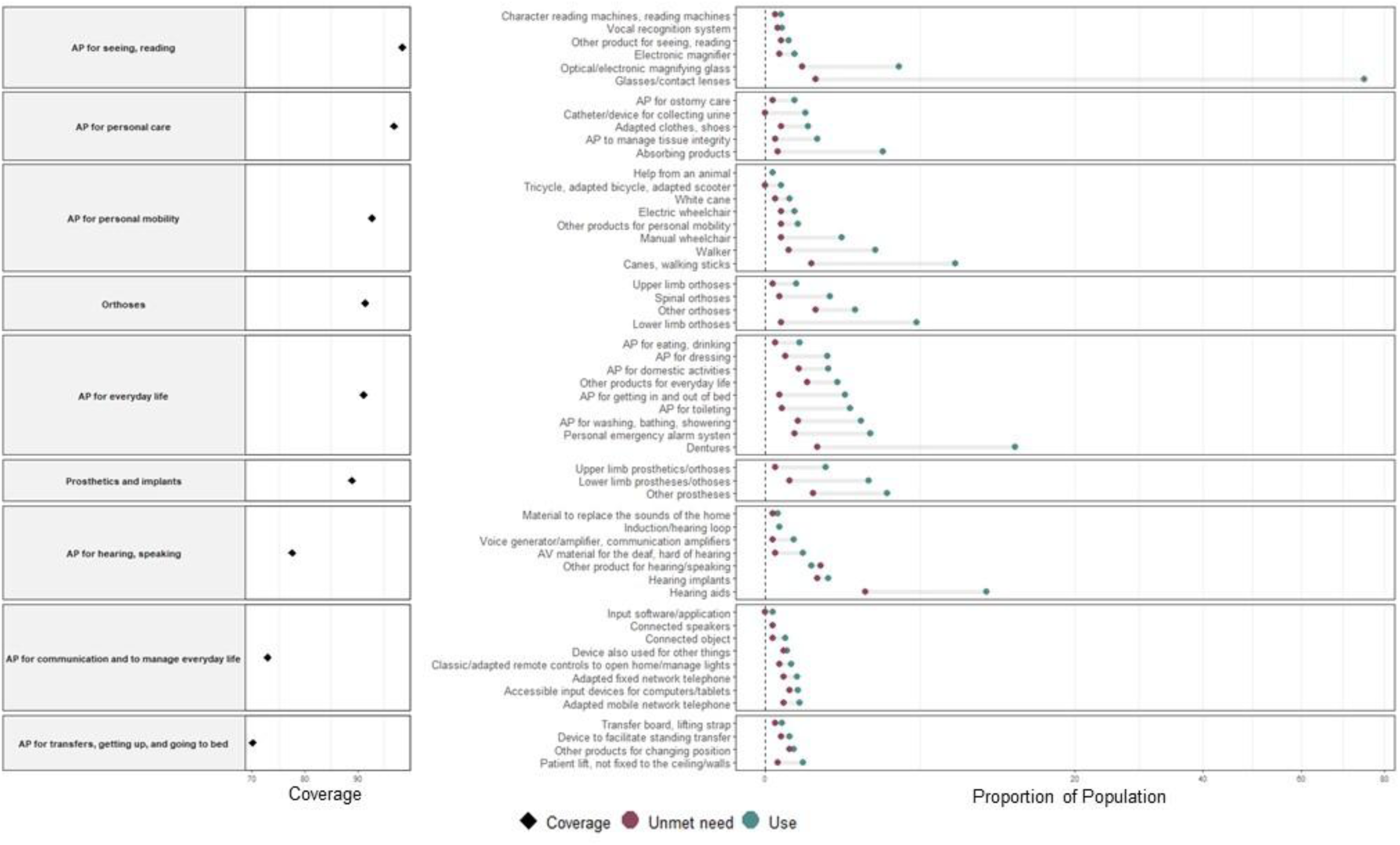
Coverage across all nine functional domains for Assistive Products (left) and Use, Unmet Need for the specific devices within each of these domains (right) Abbreviations: AP: Assistive Products Use: the proportion of the population reporting use of a particular tool – an assistive product, home modification or human help, Unmet Need: the proportion of the population who reports needing a tool but does not use one, Coverage: the proportion of individuals who use or need a tool whose need is met

Among home modifications (**Figure 3**; **Table S10**), adaptations in the bathing area were the most commonly reported (use 16·1%; coverage 70·6%), followed by modifications in the toilet areas (10·2%; 79·8%), stairs (7·9%; 81·5%), other areas (4·2%; 75·9%), and kitchen (1·1%; 59·5%). Among respondents reporting need for home modifications they did not have, most declared cost (40·3%), inability to make these modifications in their home (14·9%) and difficulties obtaining required modifications (14·6%) as the underlying reasons (**Table S12**).

**Figure 3:**
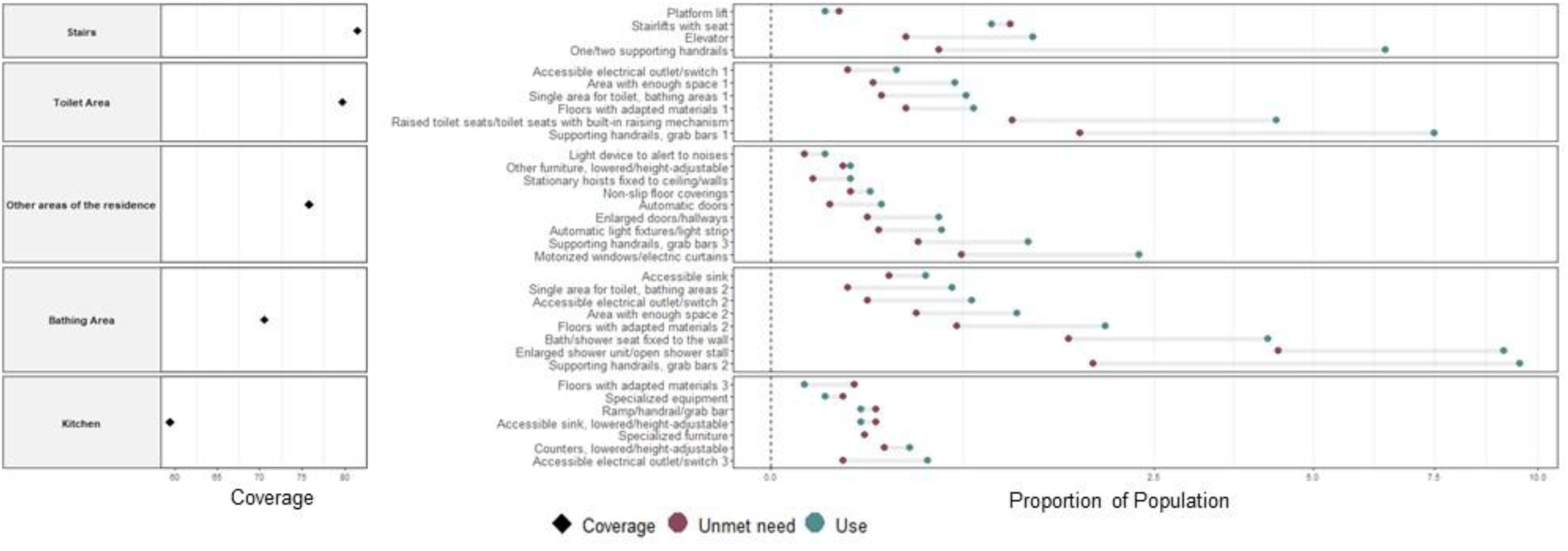
Coverage across all five areas of the home for Home Modifications (left) and Use, Unmet Need for the specific modifications within each of these areas (right) Abbreviations: HM: Home Modifications Use: the proportion of the population reporting use of a particular tool – an assistive product, home modification or human help, Unmet Need: the proportion of the population who reports needing a tool but does not use one, Coverage: the proportion of individuals with total need for a tool whose need is met

The activities most commonly requiring human help involved undertaking less frequent tasks (use 10·3%; coverage 73·5%) and shopping for groceries (use 10·1%; coverage 84·6%) (**Figure 1; Table S10)**. In contrast, the activities least frequently requiring human help were related to ADL.

### Access Indicators - Subgroups

Use (**Figure 4**, **Tables S13-S15**) and unmet needs (**Figure 5, Tables S16-S18**) were higher in all predefined vulnerable subgroups relative the total population. Among women, individuals with lower educational attainment, those with chronic conditions, and people with multimorbidity, use and unmet need were always less than twice that observed in the total population but still consistently larger than that observed in the total population (**Figures 4, 5**).

**Figure 4:**
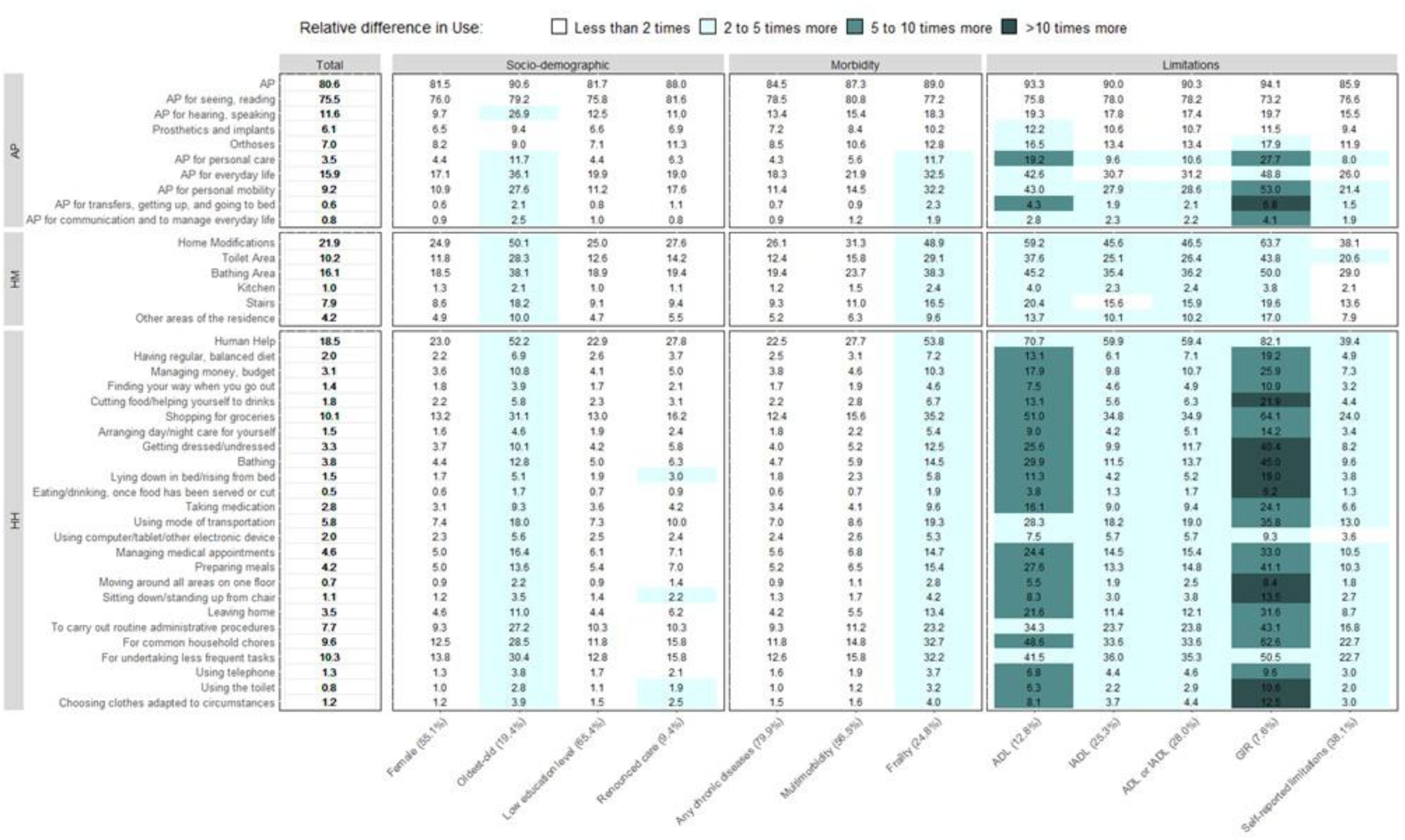
Comparing the proportion of the population that use Assistive Products, Home Modifications and Human Help in the total population to the proportion of users in vulnerable subgroups. Abbreviations: AP: Assistive Products, HM: Home Modifications, HH: Human Help For each subgroup, the proportion of members from the total population belonging to the subgroup is noted Use: the proportion of the population reporting use of a particular tool – an assistive product, home modification or human help

**Figure 5:**
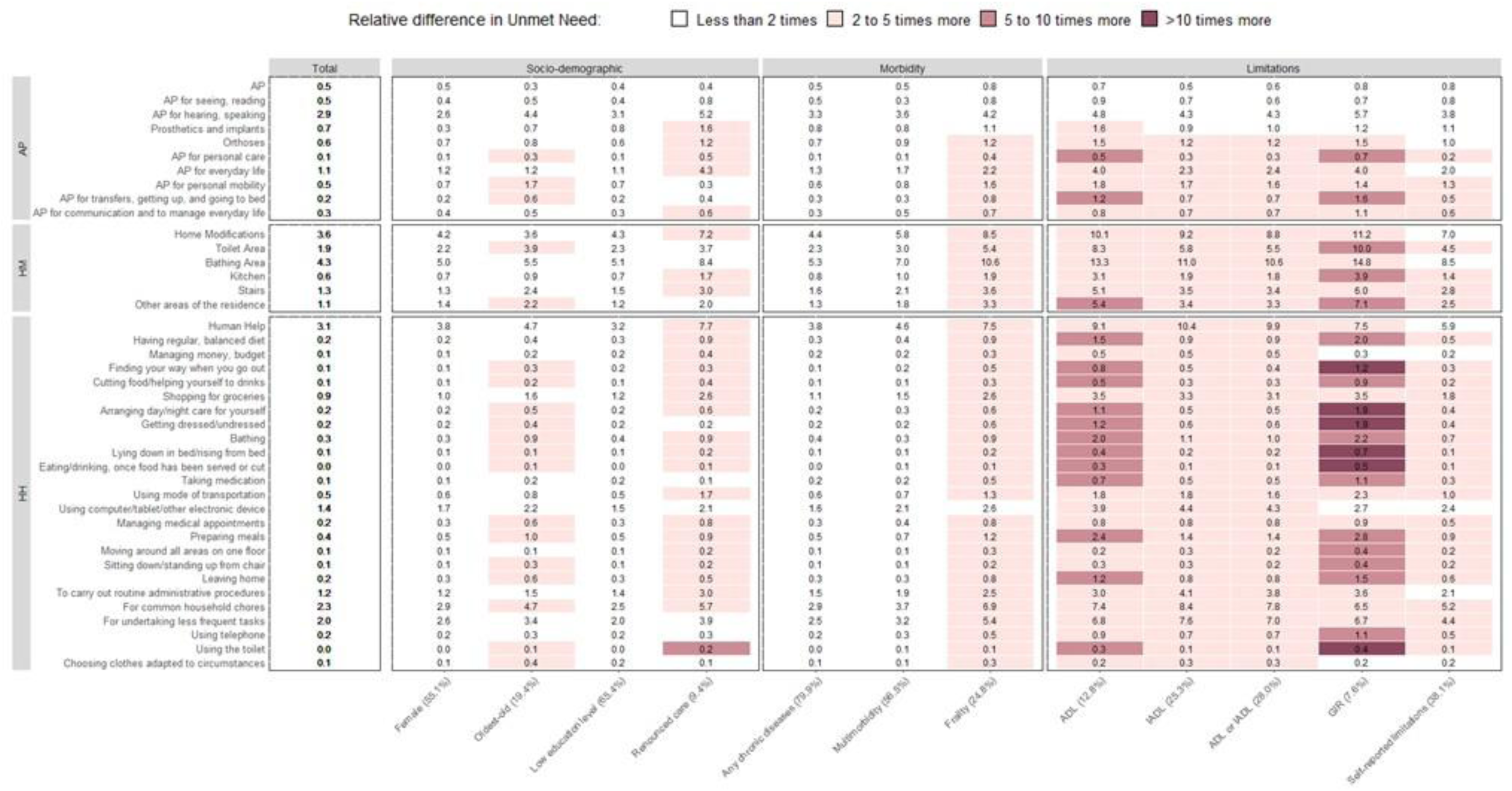
Comparing the proportion of the population that have unmet need for Assistive Products, Home Modifications and Human Help in the total population to the proportion in vulnerable subgroups. Abbreviations: AP: Assistive Products, HM: Home Modifications, HH: Human Help For each subgroup, the proportion of members from the total population belonging to the subgroup is noted Unmet Need: the proportion of the population who reports needing a tool but does not use one

Among the oldest old, use of human help, home modifications, and six of the nine functional groups for assistive products was between 2 and 5 times larger than that in the total population. Unmet needs were mostly between two and five times that observed in the total population (three of nine categories of assistive products, two of five areas of the home for home modifications, 14 of 24 human help needs). Use of assistive products among individuals who reported foregoing care during the past year was similar to that of the total population overall, although it was approximately double for four of the activities requiring human help. In contrast, unmet needs were more frequent than in the total population for a large number of assistive products, with 25 of the 38 tools showing at least a twofold higher prevalence of unmet need.

Among frail individuals, use was between two and five times higher for all categories of human help, home modifications, and most assistive product categories than that observed in the total population. The relative difference in unmet needs for this subgroup were of a similar magnitude to those observed for use and followed a comparable distribution across product categories.

Differences with the total population were more pronounced among individuals with limitations. Among those reporting limitations in ADL, 36 of the 38 tools had use and unmet need more than two times higher than in the total population. For many of these, the relative differences between those with ADL limitations and the total study population were between 5 and 10. Other definitions of disability exhibited similar use and unmet need patterns; across most categories of assistive technologies, the differences were between two and five times that in the total population. Use and unmet need were above two times greater among those in GIR groups 1 through 4 than in the total study population for all areas of the home, all activities requiring human help and six of the nine functional groups for assistive products. Notably, for nine of the activities requiring human help and for assistive products used for transfers, use was more than ten times higher for this subgroup. Similarly, unmet need for six of the activities requiring human help was more than ten times larger among those considered GIR groups 1 through 4.

## Discussion

Using data from a large, representative survey of community-dwelling adults 60 years and older, this study provides a comprehensive assessment of the use, needs, and coverage of a wide range of assistive technologies, including assistive products, home modifications, and human help. Four main findings emerge. First, 83·2% of older adults reported using at least one assistive technology, with 80·6% using an assistive product, 21·9% reporting a home modification, and 18·5% receiving human help for daily activities. When eyeglasses were excluded, the use of assistive technologies and assistive products decreased to 46·5% and 35·7%, respectively. Second, coverage (the extent to which identified needs were met by appropriate resources) ranged from 50·1% to 98·5%, with lower coverage observed for assistive products for hearing, speaking and for communication, as well as human help. Third, estimates varied markedly across subgroups, with higher use and higher unmet needs observed among participants with frailty or functional limitations, and comparable levels of use but greater unmet needs among participants reporting renunciation of care. None of the vulnerable subgroups demonstrated better access to assistive products than the total population. Finally, cost was the most frequently reported reason for renouncing an assistive product or home modification. Taken together, these findings highlight persistent barriers to accessing certain categories of assistive resources and identify specific population groups with unmet needs.

Comparison with previous studies or cross-national comparisons remain challenging due to the absence of widely accepted definitions and categorizations for assistive products, the heterogeneity of products included in different surveys, and the lack of harmonized indicators to assess accessibility.^18,23^ These methodological differences also extend to the populations studied and notably the choice of a reference population, as estimates differ greatly depending on whether they are estimated across the total population (adults ≥60 years) or restricted to older adults who report functional limitations. In the present study, indicators were calculated in the total population (adults ≥60 years) using a previously published set of access indicators to provide a population-level perspective relevant for public health monitoring and to improve comparability across studies. Subgroup analyses conducted among participants with limitations, based on three alternative measures of activity limitations, complement this approach and also illustrate how estimates may vary depending on how limitations are defined.

Population-based evidence on the use and accessibility of assistive products among older adults remains relatively scarce, despite its importance for informing policies that support healthy ageing and ageing in place, or in the comfort of one’s home. Existing studies conducted in the general older population suggest substantial use of assistive products but also highlight important variability in both the products considered and the populations studied. Estimates of assistive product use among older adults in European surveys, for example, range from 50% to 68%, depending on the scope of devices included and the age groups considered.^9^ Among studies excluding widely used devices such as eyeglasses or dentures from the definition of assistive products, hearing aids and home modifications frequently emerge as commonly used products, a pattern that is consistent with our findings.

More broadly, the evidence base remains fragmented because many studies focus on categories or specific devices rather than the full spectrum of assistive products that are likely to be relevant to ageing populations. Research on digital and connected devices illustrates this pattern. Studies among older adults have shown high use of information and communication products, such as home monitoring tools or connected health devices,^24,25^ as well as the uptake of specific devices such as connected televisions.^26^ Such studies provide valuable insights into the diffusion of emerging technologies but their design rarely allows the simultaneous assessment of use, needs, and coverage across the broader range of assistive products salient for all older adults.

This fragmentation of evidence also reflects broader structural challenges in monitoring assistive product accessibility. For example, at the national level, a recent French study^27^ emphasized the absence of integrated information systems capable of tracking assistive product provision and the derivation of indicators beyond use of such devices. This challenge is further compounded by the complexity of reimbursement mechanisms, the presence of out-of-pocket expenditures that are difficult to quantify, and the coexistence of several funding schemes linked to disability and ageing policies.^28,29^

Our study helps address these gaps by providing population-based estimates of assistive product use, need, and coverage across a wide range of products among older adults. Using nationally representative survey data, it offers an integrated perspective that complements existing studies on individual categories or devices and supports the development of population-level monitoring of assistive product access.

Beyond studies conducted in the older adult’s general population, a substantial body of research has examined assistive product use among individuals with limitations^17^ and in the total population including, but not focused on, older adults.^10^ Reviews that synthesized evidence from multiple countries report particularly large variations in the use of visual (from 1 to 87%), hearing (5 to 90%), and mobility (4 to 68%) aids.^10^ Similarly, international studies on priority assistive products identified by the WHO (hearing aids, prosthetic limbs, wheelchairs, eyeglasses, and personal digital assistants) show wide differences across settings.^11^ These studies consistently highlight substantial gaps in coverage, particularly for mobility-related devices, and indicate that unmet needs remain common even in settings with established provision of assistive products. These observations are broadly consistent with our findings, which show that both use and unmet need are considerably higher among individuals with limitations.

In the current study, frail individuals also showed higher levels of use and unmet need. These patterns are consistent with the broader literature suggesting that health status, multimorbidity, and frailty are strongly associated with both the need for, and the adoption of assistive products.^30–33^ The oldest participants and women reported higher use and higher unmet needs consistent with previous findings.^16,34^ The absolute number of individuals requiring assistive products, and potentially experiencing unmet needs, is likely to increase substantially in the coming decades as populations age and the proportion of women increases in the oldest age groups.

Cost was the most frequently reported reason for renouncing assistive products and home modifications^12,13^ and may reflect substantial out-of-pocket expenditures for certain products, particularly when reimbursement schemes are incomplete or access to them is complex. In addition to financial barriers, a large proportion of respondents selected “other” in response to the question on barriers to access, despite the availability of seven other predefined options including psychological barriers such as stigma. This suggests that barriers may be difficult to categorize and may reflect a combination of social, psychological, organizational, and informational factors.^12,14^ Importantly, the present study was not designed to investigate service delivery, although previous research indicates that the organization of assistive technology services, including assessment, prescription, training, and follow-up, plays a key role in determining effective access.^13,15^

The rapid evolution of assistive technologies also raises important questions for future health systems. Advances in digital technologies have led to the development of a growing range of connected or smart assistive devices designed to support independent living and health monitoring. Such technologies may play an increasingly important role in ageing societies, particularly in contexts where demand for human assistance is expected to rise while the availability of both professional and informal caregivers remains uncertain.^35^ In our study, the need for human assistance was substantial and its coverage appeared lower than that observed for other technologies, suggesting potential opportunities for complementary technological solutions. However, despite rapid technological innovation, relatively few emerging assistive technologies have demonstrated robust effectiveness in older populations,^36,37^ highlighting the need for further evaluation. Recent studies also suggest that the expected benefits for certain assistive tools cannot be extrapolated to all subgroups of the older population, such as frail individuals.^38^ Several limitations merit acknowledgment. First, the assessment relied on self-reported information and assumed that participants were aware of the resources relevant to their needs, whereas previous research has highlighted limited awareness of assistive technologies in the general population.^15^ Second, some important subgroups, such as institutionalized older adults or older adults with cognitive impairment, were not included in the survey, although they may experience specific needs and barriers related to assistive product use.

In conclusion, this large population-based study highlights substantial use of assistive technologies among older adults but also reveals important unmet needs, particularly for human assistance and among vulnerable subgroups such as individuals with functional limitations or frailty. These findings underscore persistent inequalities in access to assistive products that support autonomy in ageing populations. Strengthening population-level monitoring, improving financial accessibility, and integrating emerging assistive technologies in policies that promote healthy ageing policies will be critical in rapidly ageing societies.

## Author contributions

GCS and BL conducted the literature review. GCS and BL completed the analyses and worked on the preparation of figures and tables. AF provided statistical and data management support. AF, LJ, SS, ASM, helped interpret all findings. GCS and BL wrote the first draft of the manuscript, which was then reviewed and approved by all co-authors.

## Acknowledgments

This project is funded by a grant from “France 2030 ANR-23-PAVH-0006” and by IReSP/CNSA as part of the 2023 call for projects for the research support program “Older people, people with disabilities at all stages of life, relatives, and professionals,” supported by the National Solidarity Fund for Autonomy (CNSA) “AAP-2023-APAOB-318115”.

## Data Availability

Data from Autonomy (Autonomie) are available upon request by following the procedure described in https://drees.solidarites-sante.gouv.fr/sources-outils-et-enquetes/lenquete-autonomie-menages-2022-volet-individus.

