## Supplementary Material for "Access to assistive products, home modifications and human help in older adults in the 2021-2025 Autonomy survey"

**Gabriella C SILVA, PhD,[1] Aurore FAYOSSE, MSc[1] Louis JACOB, PhD[1,2,3] Séverine SABIA, PhD[1,4] Archana SINGH-MANOUX, PhD[1,4] Benjamin LANDRÉ, PhD*[1]**

[1]Université Paris Cité, Inserm U1153, CRESS, Epidemiology of Ageing and Neurodegenerative diseases, Paris, France
[2]Université Paris Cité, AP-HP, Lariboisière-Fernand Widal Hospital, Department of physical medicine and rehabilitation, Paris, France
[3]Research and Development Unit, Parc Sanitari Sant Joan de Déu, CIBERSAM, ISCIII, Dr. Antoni Pujadas, 42, Sant Boi de Llobregat, Barcelona, Spain
[4]UCL Brain Sciences, University College London, UK

**Supplementary method 1: Definition of access indicators**

Access indicators were defined based on the framework proposed by Danemayer et al. (2002) (Table 1 in the original article) and adapted to the variables available in the *Autonomy* survey.

We defined six access indicators: **(1) Use, (2) Total Need, (3) Met Need, (4) Undermet Need, (5) Unmet Need,** and **(6) Coverage**.

**Indicators available at the specific level**

Due to the design of the survey, **Met Need**, **Undermet Need**, and **Coverage** cannot generally be estimated at the level of individual AP or specific HM. With the exception of glasses, hearing aids, and wheelchairs, respondents were not asked whether the specific product or modification they used was appropriate or required replacement.

Therefore, only **Use**, **Total Need**, and **Unmet Need** can be estimated for:

- all 54 specific assistive products (AP) across the nine functional groups, and
- all 34 home modifications (HM) across the five areas of the home

**Indicators available at the group category level**

All six access indicators can be derived at the group and overall level for:

- the nine functional group levels for AP plus the overall level for AP,
- the five areas of the home for HM plus the overall level for HM, and
- the 24 activities potentially requiring HH plus the overall level for HH.

**Operational definition of met and undermet need**

To derive these indicators, we applied the following assumptions.

A participant was considered to have **met need** within a functional group (for AP) or a home area (for HM) if they reported using at least one product or modification within that group or area and did not report needing any additional product or modification within the same group or area.

A participant was considered to have **undermet need** for the functional group if they reported using an AP within a functional group but also needing another AP within the same functional group. Similarly, a participant was considered to have undermet need for HM in an area of the home if they had a home modification within that area of the home but reported needing an additional modification within the same area. In these cases, the existing device or modification was considered **insufficient to fully meet the individual's needs**.

Definitions of all indicators, with illustrative examples, are presented in **Panel A**.

**Panel A: Definitions of access indicators**

| Access indicator | Definition | Calculation |
|---|---|---|
| **At AP or HM level** | | |
| **Access indicator** | **Definition** | **Calculation** |
| **Use** of AP/HM | Proportion of the population* reporting use of a given AP or HM | (Population who Uses an AP/HM)/Population |
| *Example: Use of walker* | *Proportion of the population reporting use of a walker* | *(Population using a Walker) / Population* |
| **Total Need** of AP/HM | The proportion of the population who could benefit from using or use a specific AP/HM | (Population who Needs and/or Uses an AP/HM)/Population |
| *Example: Total Need for Walker* | *The proportion of the population who use a Walker or could benefit from one* | *(Population who Use or Need a Walker) / Population* |
| **Unmet Need** for AP/HM | The proportion of the population who do not use the AP/HM but declared needing it | (Population who Needs an AP/HM but does not use this AP/HM)/ Population |
| *Example: Unmet Need for Walker* | *The proportion of the population not using a Walker and that expressed needing it* | *(Population who Need a Walker but do not Use a Walker) / Population* |
| **At functional level for AP or home area level for HM** | | |
| **Access indicator** | **Definition** | **Calculation** |
| **Use** of AP/HM in group category | The proportion of the population that declared using any of the AP/HM within the group category | (Population who use at least one of the AP/HM within the group category)/Population |
| *Example: Use of AP for personal mobility* | *The proportion of the population that expressed using any of the AP (canes or walking sticks, white cane, ..., other mobility products) within AP for personal mobility* | *(Population who Use a 1. Cane or Walking Stick, 2. White Cane, ..., 8. Other Mobility Products)/ Population* |
| **Total need** for AP/HM group category | The proportion of the population who could benefit from using or use any of the AP/HM within the group category | (Population with Need or Use for any of the AP/HM within the group category)/ Population |
| *Example: Total Need for AP for personal mobility* | *The proportion of the population that expressed using or needing any of the AP (canes or walking sticks, white cane, ..., other mobility products) within AP for personal mobility* | *(Population who Use or Need a 1. Cane or Walking Stick, 2. White Cane, ..., 8. Other Mobility Products)/ Population* |
| **Undermet need** of AP/HM group category | The proportion of the population that uses an inappropriate AP in the functional group, defined as reporting needing a different AP within the same functional group as an AP that is used | (Population who Use an inappropriate AP within the group category) / Population |
| *Example: Undermet Need for APs for personal mobility* | *The proportion of the population that uses any of the APs for personal mobility and also report needing any of the other APs for personal mobility* | *(Population who 1. Use a Cane or Walking stuck AND need a white cane, walker, ..., or other products for personal mobility, 2. Use a White Cane AND need a cane or walking stick, walker, ..., or other products for personal mobility, ...) / Population* |

| | | |
|---|---|---|
| **Unmet need** for AP/HM group category<br>*Example: Unmet Need for APs for personal mobility* | The proportion of the population that does not use any of the APs within the functional group but that expressed needing at least one of the APs within this category<br>*The proportion of the population that does not use any APs for personal mobility but report needing an AP for personal mobility* | (Population who need an AP in the functional group but do not currently use any APs within the group category) / Population<br>*(Population not using any APs for personal mobility but need an AP for personal mobility)/ Population* |
| **Coverage** for AP/HM group category<br>*Example: coverage for AP for personal mobility* | The proportion of the population with need for an AP/HM in the functional group who are using the appropriate AP/HM within the group category<br>*Proportion of individuals who need mobility devices and whose needs are met* | Met Need for group category /<br>Total Need for group category<br>*Met need for personal mobility APs / Total need for personal mobility APs* |

**Abbreviations**: AP: Assistive products; HM: Home Modification; HH: Human Help.

*The population can be the full population of interest or a subsample of particular interest (ex. those with need)

**Supplementary method 2: deviations from the pre-registered protocol.**

The protocol was pre-registered on OpenScience Framework on March 19 2025 before the data was obtained.

It can be viewed at the following link: OSF | Accessibility to assistive devices, human help and home amendments in older adults: results from the Autonomie survey.

Deviations from the initial protocol are described in the table below.

| Original text from protocol | Modification + Reason |
|---|---|
| What are the differences in accessibility between the general population aged 60 and over and sub-groups of the population considered to be at greater risk?<br><br>In subgroups based on socio-demographic characteristics, we expect to observe worse performance on accessibility indicators among: • Women compared with Men • Those in low socioeconomic position compared with non-low socioeconomic position • People living in rural areas compared with people living in urban areas • Older participants compared with younger ones<br>Baseline comparisons between categories of a given subgroup (ex. rural vs. urban, older vs early old) will be based on p-values from the weighted chi² test and weighted confidence intervals. | We described comparisons between the subgroup considered vulnerable or at risk and its counterpart. However, due to space limitations and the already extensive length of our manuscript/analyses, we have instead decided to focus primarily on our second objective involving comparisons between the vulnerable subgroup and the general population. As a result, some of the statistical analyses involving comparisons between the categories of subgroups have not yet been completed. We hope to develop this analysis further in another manuscript. |
| A weighted logistic regression model with an indicator for non-use as the outcome and the previously mentioned set of variables as predictors will be estimated to identify which sociodemographic factors and health characteristics contribute to non-use. | Because the manuscript is extensive and there is a lot of information to report, we plan to develop this in another set of analyses. |
| For convenience, in the following paragraphs we will refer to assistive devices, home modifications and human-assisted activities as assistive products. | After reviewing the literature more thoroughly, we now refer to assistive devices as assistive products and all three (assistive devices, home modifications and human help) as assistive technology. |
| Participants who reported needing a home adaptation could also report one or more reasons for non-use among 10: (1) It costs too much, …., (9) You don't think it will be of any lasting use and (10) For another reason. | We planned to report all of the reasons for non-use of a home modification but the dataset contained some inconsistencies with the variable for reason (10), corresponding to “Other”. Therefore, we do not report the proportion of people needing a home modification that reported this reason. |
| Socio-economic position: defined using standardized education levels and categorized as low (none or primary education), intermediate (baccalaureate level) and high (university or higher degree) education levels; and, if available, defined by income and categorized as low, intermediate and high levels using categories provided by INSEE. | Although we wanted to consider income as a sociodemographic variable and define those with low-income as a subgroup of interest, we were unable to because it was unclear what exact income information we had received. For example, we were unable to determine how many people in the household were working. As a result, mapping it into meaningful categories for subgroups analyses was not possible. |
| Type of living area: defined as urban or rural based on the categorization provided by INSEE. | We did not receive information on the type of living area and were thus unable to create an urban versus rural indicator for use in subgroup analyses. |
| Frailty status: defined based on the five criteria (weakness, slowness, exhaustion, low physical activity and shrinking) of the Fried et al.2 frailty phenotype and categorized as robust | Due to the large number of subgroups already considered, for simplicity we categorized participants into frail versus not frail. The not |

| | |
|---|---|
| (0 impaired criteria), prefrail (1 or 2 impaired criteria) or frail (3 or more impaired criteria). Criteria will be adapted to the data available in the self-reported survey. | frail group therefore included the robust and prefrail participants. |
| Multimorbidity: will be defined based on the number of reported chronic diseases from a list of 37 diseases, summarized in file Table 5. | Although file Table 5 in the protocol included the correct list, the protocol should have stated 47 diseases, not 37. |
| Participants will also have the opportunity to report conditions not mentioned in the list; these will be counted if they correspond to a chronic disease. | When defining multimorbidity, we opted to only use the conditions from the list. |
| Disability: 3 categorizations of disability will be used. | We decided to include an additional indicator of disability based on whether the respondent self-reported feeling limited in everyday activities for at least 6 months. |
| Two categorizations are based on the basic activity limitations of daily living (BADL) of Katz et al. (difficulty eating, getting in/out of bed or chair, using toilet, dressing, bathing/showering, walking) and the instrumental limitations of daily living (IADL) of Lawton et al. (using telephone, doing day-to-day administrative procedures, preparing meals, shopping, doing light work (such as washing up, laundry, tidying up, cleaning, etc.), doing occasional heavy work (such as small tasks, washing tiles, etc.)). We will define the presence or absence of limitations in the BADL or the IADL. We will define the BADL and IADL stages based on the classification provided by Stineman et al. | The list of activities considered for ADL and IADL varies slightly from what is mentioned in the protocol. The list included in the protocol does not align with the list used for Stineman classifications. ADL included eating, using the toilet, dressing, bathing, getting in and out of the bed/chair, and walking; IADL included using the telephone, money management, meal preparation, light housework, heavy housework, and shopping. |
| We will define 5 categories: GIR 1, GIR 2, GIR 3, GIR 4, no autonomy aids received. The categories corresponding to the greatest loss of autonomy (GIR 1 and GIR 2) may be merged due to low numbers. | GIR groups 1 through 4 were merged for the subgroup analyses due to low counts. The interpretation is still intuitive though: GIR groups 1 through 4 are those eligible for increased financial assistance. |

**Table S1: List of 54 assistive products distributed by 9 functional domains, and 34 home modifications distributed by 5 area of the home, with original (French) and translated terms (English) and with closest ISO 9999 code.**

| Functional Domain | | Assistive product | | ISO 9999[b] |
|---|---|---|---|---|
| English[a] | French | English[a] | French | |
| 1. Assistive products for seeing, reading | Aides pour voir, lire | 1. Glasses/contact lenses | Lunettes ou lentilles | 22 03 06 |
| | | 2. Optical or electronic magnifying glass | Loupe optique ou électronique | 22 03 09 |
| | | 3. Electronic magnifier | Agrandisseur électronique | 22 03 09 |
| | | 4. Vocal recognition system | Système de reconnaissance vocale ou de synthèse vocale | 22 21 12<br>22 21 09<br>22 39 07 |
| | | 5. Character reading machines, reading machines | Système de reconnaissance de caractères, machine à lire | 22 30 21 |
| | | 6. Other product for seeing, reading | Autre objet ou aide technique pour voir | Not applicable |
| 2. Assistive products for hearing, speaking | Aides pour entendre ou parler | 7. Hearing implants | Implant auditif | 22 06 21 |
| | | 8. Hearing aids | Prothèse auditive | 22 06 06<br>22 06 09<br>22 06 12<br>22 06 15 |
| | | 9. Material to replace the sounds of the home | Matériel pour remplacer les sons de la maison | |
| | | 10. Audio visual material for the deaf and hard of hearing | Matériel audio-visuel adapté pour les malentendants | |
| | | 11. Induction loop or hearing loop | Boucle magnétique | 22 18 30 |
| | | 12. Voice generator or amplifier, Communication amplifiers | Générateur, amplificateur de voix, canule phonatoire | 22 09 03<br>22 09 06<br>22 21 06 |
| | | 13. Other product for hearing or speaking | Autre objet ou aide technique pour communiquer | Not applicable |
| 3. Prosthetics and implants | Prothèses et implants | 14. Upper limb prosthetics or orthoses (finger, hand or arm, etc.) | Prothèse externe ou orthèse des membres supérieurs (doigt, main ou bras artificiel, etc.) | 06 18 |
| | | 15. Lower limb prostheses or orthoses (foot or leg, etc.) | Prothèse externe ou orthèse des membres inférieurs (pied ou jambe artificielle, etc.) | 06 24 |
| | | 16. Other prostheses (eye, nose, etc.) | Autre prothèse (oeil de verre, prothèse mammaire, prothèse de nez, etc.) | 06 30 |

| | | | | |
|---|---|---|---|---|
| 4. Orthoses | Appareillage de soutien, de maintien ou de correction de la position du corps | 17. Lower limb orthoses (orthopedic socks or insoles, etc.) | Appareillage des membres inférieurs (chaussures ou semelles orthopédiques, etc.) | 06 12 |
| | | 18. Upper limb orthoses | Appareillage des membres supérieurs | 06 06 |
| | | 19. Spinal orthoses (corset, etc.) | Appareillage du tronc ou de la colonne vertébrale (corset, etc.) | 06 03 |
| | | 20. Other orthoses | Autre appareillage de soutien, de maintien ou de correction de la position du corps | Not applicable |
| 5. Assistive products for personal care | Aides pour les soins personnels | 21. Catheter or device for collecting urine | Sonde ou collecteur d'urines (ou autre système d'évacuation de l'urine) | 09 24<br>09 27 |
| | | 22. Absorbing products | Protections absorbantes (couches) | 09 30 18<br>09 30 21 |
| | | 23. Adapted clothes and shoes | Vêtements adaptés | 09 03 |
| | | 24. Assistive products for ostomy care | Aides pour stomisés (poches, absorbants) | 09 18 |
| | | 25. Assistive products intended to manage tissue integrity | Matériel anti-escarres : coussins, matelas, etc. | 04 33 |
| 6. Assistive products for everyday life | Aides à la vie quotidienne | 26. Dentures | Dentier | 06 30 36 |
| | | 27. Assistive products for washing, bathing and showering | Aides pour vous laver : brosse à dos adaptée, siège de douche non fixé au mur, planche de baignoire, etc. | 09 33 |
| | | 28. Assistive products for dressing | Aides pour vous habiller : enfile-bas, chausse-pieds à long manche, crochet à boutons, etc. | 09 09 |
| | | 29. Assistive products for eating and drinking | Aides pour manger et boire : couverts coudés, gobelet à bec, gobelet à paille, assiette à rebord, assiette compartimentée, etc. | 15 09 |
| | | 30. Assistive products for toileting | Aides pour aller aux toilettes : rehausseur non fixé à la cuvette, matériel pour faciliter l'hygiène intime, chaise percée, etc. | 09 12 |
| | | 31. Assistive products for domestic activities: system to recognize money or pay, system to help in the use of kitchen utensils, etc. | Aides pour faire les courses, préparer le repas, entretenir votre logement : système pour reconnaître l'argent ou pour payer, système pour aider dans l'utilisation des ustensiles de cuisine, etc. | 15 03<br>15 06<br>15 12<br>15 15 |
| | | 32. Assistive products for getting in and out of bed: hospital bed, electrical bed | Aides pour se lever ou se coucher : lit médicalisé, lit électrique (hors lit médicalisé) | 18 12 |
| | | 33. Personal emergency alarm system | Téléalarme | 22 27 18 |
| | | 34. Other products for everyday life | Autres aides pour les soins ou la vie quotidienne | Not applicable |

| | | | | | |
|---|---|---|---|---|---|
| 7. Assistive products for personal mobility | Aides au déplacement | 35. | Canes or walking sticks | Cannes ou béquilles | 12 03 03 |
| | | 36. | White cane | Canne blanche | 12 39 03 |
| | | 37. | Walker | Déambulateur | 12 06 |
| | | 38. | Manual wheelchair | Fauteuil roulant manuel | 12 22 |
| | | 39. | Electric wheelchair | Fauteuil roulant électrique | 12 23 |
| | | 40. | Tricycle, adapted bicycle, adapted scooter | Tricycle, vélo adapté, trottinette adaptée | 12 23 03 |
| | | 41. | Help from an animal such as a guide dog | Aide animalière comme un chien guide | Not included |
| | | 42. | Other products for personal mobility | Autres aides pour marcher ou se déplacer | Not applicable |
| 8. Assistive products for transfers, getting up, and going to bed | Aides pour les transferts, se lever et se coucher | 43. | Patient lift, not fixed to the ceiling or walls | Un lève-personne, non fixé au plafond ou aux murs | 12 36 |
| | | 44. | Transfer board or lifting strap | Une planche de transfert ou sangle de transfert | 12 31 03 |
| | | 45. | A device to facilitate standing transfer | Un matériel pour faciliter le transfert debout (guidon de transfert pivotant ou non, etc.) | 12 31<br>12 36 |
| | | 46. | Other products for changing position | Autres aides pour les transferts | Not applicable |
| 9. Assistive products for communication and to manage everyday life | Aides pour communiquer/gérer actes de la vie quotidienne | 47. | Accessible input devices for computers or tablets | Ordinateur ou tablette adapté (exemple : caractères plus gros, clavier ou souris adapté, utilisation simplifiée, volume du son élevé, fonctionnalités supplémentaires, etc.) | 22 36 |
| | | 48. | Adapted fixed network telephone | Téléphone fixe adapté | 22 24 03 |
| | | 49. | Adapted mobile network telephone | Téléphone portable adapté | 22 24 06 |
| | | 50. | Connected object (excluding speakers) | Object connecté (hors enceinte) | |
| | | 51. | Connected speakers (personal assistant) | Enceinte connectée adaptée ou non (assistant personnel) | |
| | | 52. | Input software or application | Logiciel ou application spécifique | 22 36 18 |
| | | 53. | Classic or adapted remote controls to open the home or to manage the lights | Télécommandes classiques ou adaptées pour gérer les ouvertures du logement ou les lumières | 18 21 |
| | | 54. | Device also used for other things (ex. remote for wheelchair, etc.) | Appareil aussi utilisé pour d'autres choses (commande du fauteuil roulant, etc.) | Not applicable |
| **Area of home** | | **Home modification** | | | **ISO 9999[b]** |
| **English[a]** | **French** | **English[a]** | | **French** | |
| 1. Toilet Area | Dans les toilettes | 1. | Supporting handrails and grab bars | Une ou plusieurs barres d'appui | 18 18 |

| | | | | |
|---|---|---|---|---|
| | | 2. Raised toilet seats or toilet seats with built-in raising mechanism (exclude removable booster seats) | Des toilettes surélevées ou à hauteur variable (exclure les rehausseurs amovibles) | 09 12 18<br>09 12 21 |
| | | 3. Floors with adapted materials (ex. non-slip) | Un sol spécial (par exemple anti-dérapant) | 18 24 15<br>18 33 03 |
| | | 4. Accessible electrical outlet or switch | Une prise électrique ou un interrupteur facile à utiliser | 24 09 |
| | | 5. An area with enough space (for example, for a wheelchair) | Une pièce avec suffisamment d'espace, par exemple pour un fauteuil roulant | Not applicable |
| | | 6. A single area/space for the toilet and bathing areas | Une seule pièce pour la salle de bain et les toilettes (regroupement des pièces) | Not applicable |
| 2. Bathing Area | Dans la salle de bains | 7. Supporting handrails and grab bars | Une ou plusieurs barres d'appui | 18 18 |
| | | 8. Enlarged shower unit or open shower stall (ex. Italian shower) | Une douche élargie ou avec un bac à douche au niveau du sol (douche à l'italienne par exemple) | 09 33 09 |
| | | 9. Bath/shower seat fixed to the wall | Un siège de douche fixé au mur | 09 33 05 |
| | | 10. Accessible sink (ex. height-adjustable) | Un lavabo adapté (par exemple réglable en hauteur) | 18 24 03<br>15 06 03 |
| | | 11. Floors with adapted materials (ex. non-slip) | Un sol spécial (par exemple anti-dérapant) | 18 24 15<br>18 33 03 |
| | | 12. Accessible electrical outlet or switch | Une prise électrique ou un interrupteur facile à utiliser | 24 09 |
| | | 13. An area with enough space (for example, for a wheelchair) | Une pièce avec suffisamment d'espace, par exemple pour un fauteuil roulant | Not applicable |
| | | 14. A single area/space for the toilet and bathing areas | Une seule pièce pour la salle de bain et les toilettes (regroupement des pièces) | Not applicable |
| 3. Kitchen | Dans la cuisine | 15. Counters, lowered or height-adjustable | Un plan de travail abaissé ou réglable en hauteur | 18 24 21 |
| | | 16. Accessible sink, lowered or height-adjustable | Un évier abaissé ou réglable en hauteur | 15 06 03 |
| | | 17. Specialized furniture | Des meubles spéciaux | 18 09<br>18 36 |
| | | 18. Specialized equipment (ex. for the oven or hood) | Des équipements spéciaux pour le four ou la hotte par exemple | 15 03 18<br>15 03 21<br>15 03 24 |
| | | 19. A ramp, handrail or grab bar | Une rampe ou une barre pour se tenir | 18 18 |
| | | 20. Floors with adapted materials (ex. non-slip) | Un sol spécial (par exemple anti-dérapant) | 18 24 15<br>18 33 03 |

| | | | | |
|---|---|---|---|---|
| | | 21. Accessible electrical outlet or switch | Une prise électrique ou un interrupteur facile à utiliser | 24 09 |
| 4. Stairs | Dans l'escalier | 22. One or two supporting handrails | Une ou deux rampes pour vous tenir | 18 18 |
| | | 23. Stairlifts with seat | Un siège monte-escalier | 18 30 10 |
| | | 24. Platform lift | Une plate-forme élévatrice ou un monte-charge | 18 30 05<br>18 30 08 |
| | | 25. Elevator | Un ascenseur | 18 30 03 |
| 5. Other areas of the residence | Dans le reste du logement | 26. Stationary hoists fixed to ceiling or walls | Un lève-personne fixé au plafond ou au mur | 12 36 12 |
| | | 27. Supporting handrails and grab bars | Des barres d'appui ou des rampes ailleurs dans votre logement | 18 18 |
| | | 28. Automatic light fixtures (motion-detected or remote-controlled) or light strip | Des chemins lumineux ou des interrupteurs automatiques (à détecteur de mouvement ou à télécommande) | 18 06<br>24 09 18 |
| | | 29. A light device to alert to noises (ex. by flashing) | Un dispositif lumineux pour alerter des bruits (ex : par flash) | 22 27<br>22 27 04 |
| | | 30. Motorized windows or electric curtains | Des fenêtres motorisées ou des volets roulants électriques | 18 21 06<br>18 21 09 |
| | | 31. Automatic doors (motion-detected or remote-controlled) | Des portes automatiques (à détecteur de mouvement ou à télécommande) | 18 21 03 |
| | | 32. Enlarged doors or hallways | Des portes ou des couloirs spécialement élargis | 18 24 09 |
| | | 33. Non-slip floor coverings | Des revêtements de sol anti-dérapants ailleurs dans votre logement | 18 24 15<br>18 33 03 |
| | | 34. Other furniture, lowered or height-adjustable | Des meubles abaissés ou réglables en hauteur ailleurs dans votre logement | 18 15<br>18 09<br>18 36 |

a. Autonomy is a survey in French. We translate all functional domains and assistive products, home modifications to English to the best of our capacity.

b. Approximate ISO 9999 codes (from ISO 9999 6th edition, 2016 [English]) are provided for reference. For some assistive products in the survey, an exact correspondence to one ISO 9999 is not possible or may not be applicable. We attempt to find the ISO 9999 codes that most closely match each assistive product or home modification in the survey.

**Table S2: List of 24 activities that may require human help from family members or professionals in the translated (English) and original (French) terms.**

| English | French |
|---|---|
| 1. Bathing | Pour vous laver |
| 2. Getting dressed/undressed | Pour vous habiller ou vous déshabiller |
| 3. Cutting food or helping yourself to drinks | Pour couper votre nourriture ou vous servir à boire |
| 4. Eating or drinking, once food has been served or cut | Pour manger ou boire, une fois que la nourriture est servie ou découpée |
| 5. Using the toilet | Pour vous server des toilettes |
| 6. Lying down in bed/rising from bed | Pour vous coucher ou vous lever de votre lit |
| 7. Sitting down/standing up from chair | Pour vous asseoir ou vous lever d'un siège |
| 8. Shopping for groceries | Pour faire vos courses |
| 9. Preparing meals | Pour preparer vos repas |
| 10. For common household chores | Pour les tâches ménagères courantes dans votre domicile |
| 11. For undertaking less frequent tasks | Pour les taches plus occasionnelles |
| 12. To carry out routine administrative procedures | Pour faire les démarches administratives courantes |
| 13. Taking medication | Pour prendre vos medicaments |
| 14. Moving around all the areas on one floor | Pour vous déplacer dans toutes les pièces d'un étage |
| 15. Leaving the home | Pour sortir de votre logement |
| 16. Using a mode of transportation | Pour utiliser un moyen de déplacement |
| 17. Finding your way when you go out | Pour trouver votre chemin lorsque vous sortez |
| 18. Using a telephone | Pour vous server d'un téléphone |
| 19. Using a computer, tablet or other electronic device | Pour vous server d'un ordinateur, d'une tablette numérique ou d'un objet connecté |
| 20. Choosing clothes adapted to circumstances | Pour vous choisir des vêtements adaptés aux circonstances |
| 21. Having a regular, balanced diet | Pour avoir une alimentation régulière et équilibrée |
| 22. Managing money and budget | Pour gérer votre argent, votre budget |
| 23. Managing medical appointments | Pour gérer vos rendez-vous médicaux |
| 24. Arranging day or night care for yourself | Pour vous assurer une garde de jour ou une garde de nuit |

**Table S3: List of reasons for not having an assistive product or home modification considered needed, with translated (English) and original (French) terms.**

**Assistive Product**

| English | French |
|---|---|
| 1. It is too expensive | Ça coute trop cher |
| 2. It is hard to find, obtain (availability, administrative hurdles, lack of information) | C’est difficile à trouver, à obtenir (disponibilité, démarches administratives, manque d’information) |
| 3. It is too constraining, difficult to use | C’est trop contraignant, trop difficile à utiliser |
| 4. These products are for people whose health is in a worse state than yours | C’est fait pour des personnes dont l’état de santé est plus grave que le vôtre |
| 5. You hadn’t thought about it | Vous n’y avez pas pensé |
| 6. You prefer not using one | Vous préférez vous débrouiller sans aide technique |
| 7. Using one made your health problems too obvious | Cela rendrait trop visible vos problèmes de santé/votre handicap |
| 8. You don’t believe this will help in the long-run | Vous ne pensez pas que cela serve durablement |
| 9. Other reason | Autre raison |

**Home Modifications**

| English | French |
|---|---|
| 1. It is too expensive | Ça coute trop cher |
| 2. It is hard to obtain (availability, administrative hurdles, lack of information) | C’est difficile à obtenir (disponibilité, démarches administratives, manque d’information) |
| 3. It is too constraining, difficult to use | C’est trop contraignant, trop difficile à utiliser |
| 4. These products are for people whose health is in a worse state than yours | C’est fait pour des personnes dont l’état de santé est plus grave que le vôtre |
| 5. You hadn’t thought about it | Vous n’y avez pas pensé |
| 6. You are scared of damaging your home | Vous avez peur d’abîmer/de dévaloriser votre logement |
| 7. The people you live with don’t want these | Les personnes qui habitent avec vous ne le souhaitent pas |
| 8. It is not possible to make these modifications in your current home | Il est impossible de faire ces aménagements dans votre logement actuel |
| 9. You don’t believe this will help in the long-run | Vous ne pensez pas que cela serve durablement |
| 10. Other reason | Pour une autre raison |

**Table S4: List of 10 reasons for non-use of healthcare services, with original (French) and translated (English) terms.**

| English | French |
|---|---|
| 1. You couldn't afford it (too expensive or poorly reimbursed by your health insurance) | Vous ne pouviez pas vous le payer (trop cher ou mal remboursé par votre assurance maladie) |
| 2. The waiting time for an appointment was too long | Le temps d'attente pour obtenir un rendez-vous était trop long |
| 3. You didn't have a referral letter | Vous n'aviez pas de lettre de recommandation |
| 4. You didn't have time (because of your work, family, etc.) | Vous n'aviez pas le temps (à cause de votre travail, de votre famille, etc |
| 5. You couldn't get there (transportation or access difficulties, etc.) | Vous ne pouviez pas vous y rendre (difficultés de transport, d'accès, etc |
| 6. You were scared | Cela vous a fait peur |
| 7. You wanted to wait and see if it would resolve itself | Vous vouliez attendre et voir si cela passerait tout seul |
| 8. You didn't know a good healthcare professional | Vous ne connaissiez pas de bon professionnel |
| 9. The health professional was not welcoming | Le professionnel n'était pas accueillant |
| 10. Other reasons | Pour d'autres raisons |

**Table S5: List of 47 chronic conditions considered in the survey by group of conditions.**

| | |
|---|---|
| **Cardiovascular diseases** | |
| 1. | Myocardial infarction (heart attack) or sequelae of infarction |
| 2. | Coronary artery disease, angina pectoris |
| 3. | High blood pressure |
| 4. | Stroke, cerebral attack (cerebral hemorrhage, cerebral thrombosis) or sequelae of stroke or cerebral attack |
| 5. | Heart failure |
| 6. | Arteritis of the lower limbs (arterial disease) |
| 7. | Rhythm disorders |
| 8. | Other cardiovascular diseases or problems |
| **Respiratory diseases** | |
| 9. | Asthma (including allergic asthma) |
| 10. | Chronic bronchitis, chronic obstructive pulmonary disease (COPD), emphysema |
| 11. | Allergies (excluding allergic asthma) such as allergic rhinitis, hay fever, allergic conjunctivitis, skin allergies, food allergies |
| 12. | Other respiratory diseases |
| **Diseases affecting the bones and joints** | |
| 13. | Chronic pain or disorders of the back or lower back, such as low back pain, lumbago, sciatica |
| 14. | Chronic pain or disorders of the neck or cervical spine |
| 15. | Rheumatoid arthritis |
| 16. | Other types of arthritis (joint inflammation) |
| 17. | Knee osteoarthritis |
| 18. | Hip osteoarthritis |
| 19. | Osteoarthritis in other locations |
| 20. | Osteoporosis |
| 21. | Other diseases or problems affecting the bones and joints |
| **Neurological diseases** | |
| 22. | Migraines, severe headaches |
| 23. | Epilepsy |
| 24. | Alzheimer's disease and other similar diseases |
| 25. | Parkinson's disease |
| 26. | Multiple sclerosis |
| 27. | Other neurological diseases or problems |
| **Chronic mental illnesses** | |
| 28. | Anxiety |
| 29. | Depression |
| 30. | Neurodevelopmental disorders and/or autism spectrum disorders |
| 31. | Schizophrenia |
| 32. | Bipolar disorders |
| 33. | Anorexia nervosa |
| 34. | Other mental illnesses or problems |
| **Chronic eye or ear diseases** | |
| 35. | Unoperated cataracts |
| 36. | Glaucoma with sequelae |
| 37. | Age-related macular degeneration |
| 38. | Other eye diseases |
| 39. | Ear diseases |
| 40. | Other hearing diseases or problems |
| **Other diseases** | |
| 41. | Kidney failure |
| 42. | Cirrhosis of the liver |
| 43. | Diabetes |
| 44. | Thyroid disorder (goiter, hyperthyroidism, or hypothyroidism) |
| 45. | Cancer (all malignant tumors including leukemia and lymphoma) |
| 46. | Injury or permanent sequelae caused by an accident |
| 47. | Other diseases |

**Table S6: List of activities of daily living (ADL) and instrumental activities of daily living (IADL) with survey questions used to assess each**

| | | ***Autonomy* question [English]** | ***Autonomy* question [French]** |
|---|---|---|---|
| ADL | Eating | Do you have difficulties eating or drinking alone, once the food is served and cut? | Avez-vous des difficultés pour manger ou boire seul(e), une fois que la nourriture est servie et découpée? |
| | Using the toilet | Do you have difficulties using the toilet alone? | Avez-vous des difficultés pour vous servir seul(e) des toilettes? |
| | Dressing | Do you have difficulties getting dressed or undressed alone? | Avez-vous des difficultés pour vous habiller ou vous déshabiller seul(e)? |
| | Bathing | Do you have difficulties bathing alone? | Avez-vous des difficultés pour vous laver seul(e)? |
| | Getting in/out of bed/chair | Do you have difficulties getting up or laying down alone in your bed?<br>Do you have difficulties sitting down or rising from a chair? | Avez-vous des difficultés pour vous coucher ou vous lever seul(e) de votre lit?<br>Avez-vous des difficultés pour vous asseoir ou vous lever seul(e) d'un siège? |
| | Walking | Do you have difficulties moving around all the spaces of a floor without someone's help? | Avez-vous des difficultés pour vous déplacer sans l'aide de quelqu'un dans toutes les pièces d'un étage? |
| IADL | Using the telephone | Do you have difficulties using a phone? | Avez-vous des difficultés pour vous servir d'un téléphone? |
| | Money management | Do you have difficulties managing your money and budget without help? | Avez-vous des difficultés à gérer sans aide votre argent, votre budget? |
| | Meal preparation | Do you have difficulties preparing meals without help? | Avez-vous des difficultés pour préparer vos repas sans aide? |
| | Light housework | Do you have difficulties completing common household tasks such as doing the dishes, laundry, organizing and cleaning, without help? | Avez-vous des difficultés pour faire sans aide les tâches ménagères courantes dans votre domicile telles que la vaisselle, la lessive, le rangement, le ménage…? |
| | Heavy housework | Do you have difficulties completing less frequent household tasks like cleaning the floor without help? | Avez-vous des difficultés pour faire des tâches plus occasionnelles sans aide (petits travaux, laver les carreaux, etc.)? |
| | Shopping | Do you have difficulties grocery shopping without help? | Avez-vous des difficultés pour faire vos courses sans aide? |

**Table S7: Characteristics of the survey and study population.**

| | | Weighted Percentage of Population Ages 60 and above (Observed Frequency) (number of survey respondents = 8,024; estimated population size = 16,715,451) |
|---|---|---|
| **Age,** *wMean (wSD)* | | 71.7 (0.13) |
| **Age [categorical]** | | |
| | Early old age (< 80 years old) | 80.6 (5,685) |
| | Oldest-old (≥ 80 years old)* | 19.4 (2,339) |
| **Sex** | | |
| | Male | 44.9 (3,561) |
| | Female* | 55.1 (4,463) |
| **Education**[a] | | |
| | High | 23.4 (1,341) |
| | Intermediate | 11.2 (808) |
| | Low* | 65.4 (5,826) |
| **Access to medical care** | | |
| | Did not renounce | 90.6 (6,971) |
| | Renounced* | 9.4 (1,053) |
| **Frailty status** | | |
| | Robust | 30.9 (1,219) |
| | Prefrail | 44.3 (2,756) |
| | Frail* | 24.8 (4,049) |
| **Chronic diseases** | | |
| | None | 20.1 (784) |
| | At least one* | 79.9 (7,240) |
| **Multimorbidity** | | |
| | Not multimorbid | 43.5 (2,150) |
| | Multimorbid* | 56.5 (5,874) |
| **GIR** | | |
| | Completely autonomous (Group 6) | 87.2 (5,509) |
| | Needs some help but mostly autonomous (Group 5) | 5.2 (836) |
| | Requires considerable or complete help (Groups 1-4) * | 7.6 (1,679) |
| **ADL** | | |
| | No limitation in ADL – Stineman Stage 0 | 87.2 (5,526) |
| | Mild – Stineman Stage I* | 3.0 (483) |
| | Moderate – Stineman Stage II* | 7.1 (1,390) |
| | Severe – Stineman Stage III * | 2.1 (492) |
| | Complete – Stineman Stage IV* | 0.6 (133) |
| **IADL** | | |
| | No limitation in IADL – Stineman Stage 0 | 74.7 (4,207) |
| | Mild – Stineman Stage I* | 6.1 (646) |
| | Moderate – Stineman Stage II* | 12.7 (2,111) |
| | Severe – Stineman Stage III * | 6.0 (955) |
| | Complete – Stineman Stage IV* | 0.4 (105) |
| **ADL/IADL** | | |
| | No limitations in ADL or IADL | 72.0 (3,748) |
| | At least one limitation in ADL or IADL* | 28.0 (4,276) |
| **Limited in everyday activities (self-reported)** | | |
| | No | 61.9 (2,817) |
| | Yes, but not heavily* | 22.6 (2,272) |
| | Yes, heavily limited* | 15.5 (2,935) |

**Abbreviations:** WMean: weighted mean; wSD: weighted standard deviation; GIR: Groupe Isso-Resources, a French measure of autonomy; ADL: activities of daily living; IADL: instrumental activities of daily living

Statistics are weighted percentage for target population (observed frequency among survey respondents) unless specified otherwise.

* denotes that this subcategory is (or is part of) the vulnerable subgroup for this variable

[a] Education data was missing for 49 respondents. These individuals were removed for the subgroup analysis based on education.

**Table S8: Reasons for having renounced medical care**

| | **Weighted Percentage of Population Ages 60 and above (Observed Frequency)** |
|---|---|
| **Renounced medical care** | 9.4 (1,053) |
| **Reasons** | |
| You could not afford it | 32.6 (305) |
| The waiting time for an appointment was too long | 13.5 (144) |
| You didn't have a referral letter | 0.2 (3) |
| You didn't have time | 5.0 (36) |
| You couldn't get there | 6.9 (103) |
| You were scared | 7.5 (80) |
| You wanted to wait and see if it would resolve itself | 3.6 (32) |
| You didn't know a good healthcare professional | 2.4 (28) |
| The health professional was not welcoming | 0.5 (5) |
| Other reasons | 27.9 (315) |

Statistics are weighted percentage for target population (observed frequency among survey respondents)

The weighted percentages under "Reasons" express the estimated proportion of people selecting each reason among those who declared renouncing medical care. Participants were able to select multiple reasons in this list.

**Table S9: Prevalence of chronic conditions among those reporting at least one chronic disease**

| Group of Conditions | Weighted Prevalence (Observed Frequency) |
|---|---|
| Cardiovascular diseases or problems | 36.9 (3,688) |
| Respiratory diseases | 15.6 (1,737) |
| Diseases or problems concerning the bones and joints | 47.0 (4,801) |
| Neurological problems or disorders | 8.6 (1,271) |
| Psychic or mental illnesses or disorders | 11.8 (1,577) |
| Eye or hearing diseases or problems | 23.6 (2,566) |
| Other health diseases or problems | 33.5 (3,510) |

Statistics are weighted percentage for target population (observed frequency among survey respondents) unless specified otherwise.

The weighted values express the estimated proportion of people ages 60 and above with a condition within each group of conditions.

**Table S10: Access indicators for assistive products, home modifications and human help at device or group category levels in participants aged 60 or older. Estimated proportion (95% CI) is reported.**

| | Use | Met Need | Undermet Need | Unmet Need | Total Need | Coverage |
|---|---|---|---|---|---|---|
| **Assistive Technologies (AP + HM + HH)** | 83.20 (81.77, 84.54) | 64.69 (63.09, 66.26) | 18.51 (17.43, 19.64) | 0.58 (0.38, 0.90) | 83.79 (82.37, 85.11) | 77.21 (75.86, 78.50) |
| **Assistive Technologies *[no glasses/contacts]*** | 46.46 (44.84, 48.10) | 31.51 (30.02, 33.03) | 14.96 (14.04, 15.92) | 4.14 (3.50, 4.89) | 50.60 (48.94, 52.27) | 62.26 (60.27, 64.21) |
| **AP** | 80.62 (79.14, 82.01) | 73.40 (71.84, 74.90) | 7.22 (6.49, 8.02) | 0.52 (0.33, 0.84) | 81.14 (79.68, 82.52) | 90.46 (89.44, 91.39) |
| **AP *[no glasses/contacts]*** | 35.66 (34.18, 37.17) | 31.58 (30.15, 33.04) | 4.09 (3.59, 4.64) | 3.66 (3.09, 4.33) | 39.32 (37.77, 40.88) | 80.31 (78.37, 82.11) |
| **AP for seeing, reading** | 75.52 (74.00, 76.99) | 74.90 (73.36, 76.38) | 0.62 (0.39, 1.00) | 0.49 (0.29, 0.81) | 76.01 (74.49, 77.46) | 98.54 (97.94, 98.97) |
| Electronic magnifier | 0.18 (0.11, 0.28) | | | 0.04 (0.02, 0.08) | 0.21 (0.14, 0.32) | |
| Other product for seeing, reading | 0.11 (0.05, 0.26) | | | 0.05 (0.03, 0.09) | 0.16 (0.09, 0.30) | |
| Character reading machines, reading machines | 0.05 (0.03, 0.09) | | | 0.02 (0.01, 0.06) | 0.07 (0.04, 0.12) | |
| Optical/electronic magnifying glass | 3.71 (3.18, 4.33) | | | 0.28 (0.13, 0.58) | 3.99 (3.42, 4.64) | |
| Glasses/contact lenses | 75.03 (73.50, 76.50) | 56.79 (55.12, 58.45) | 17.51 (16.33, 18.75) | 0.52 (0.31, 0.88) | 75.55 (74.03, 77.01) | 75.17 (73.54, 76.73) |
| Vocal recognition system | 0.06 (0.03, 0.10) | | | 0.03 (0.01, 0.07) | 0.09 (0.05, 0.14) | |
| **AP for hearing, speaking** | 11.59 (10.64, 12.60) | 11.29 (10.35, 12.29) | 0.30 (0.17, 0.53) | 2.94 (2.49, 3.47) | 14.53 (13.49, 15.64) | 77.66 (74.31, 80.69) |
| Voice generator/amplifier, communication amplifiers | 0.17 (0.08, 0.35) | | | 0.01 (0.00, 0.03) | 0.18 (0.09, 0.35) | |
| Other product for hearing/speaking | 0.45 (0.29, 0.69) | | | 0.64 (0.44, 0.94) | 1.09 (0.82, 1.45) | |
| Induction/hearing loop | 0.04 (0.01, 0.11) | | | NA | 0.04 (0.01, 0.11) | |
| Hearing implants | 0.82 (0.58, 1.16) | 0.70 (0.47, 1.03) | 0.11 (0.05, 0.25) | 0.57 (0.38, 0.85) | 1.39 (1.07, 1.81) | 50.12 (37.00, 63.22) |
| AV material for the deaf, hard of hearing | 0.30 (0.17, 0.52) | | | 0.02 (0.01, 0.06) | 0.32 (0.19, 0.54) | |
| Hearing aids | 10.22 (9.34, 11.18) | 8.73 (7.90, 9.64) | 1.33 (1.05, 1.70) | 2.08 (1.72, 2.51) | 12.30 (11.34, 13.33) | 70.99 (67.16, 74.54) |
| Material to replace the sounds of the home | 0.03 (0.01, 0.05) | | | 0.01 (0.00, 0.07) | 0.04 (0.02, 0.08) | |

| | Use | Met Need | Undermet Need | Unmet Need | Total Need | Coverage |
|---|---|---|---|---|---|---|
| **Prosthetics and implants** | 6.08 (5.43, 6.81) | 6.02 (5.37, 6.74) | 0.06 (0.02, 0.16) | 0.69 (0.49, 0.98) | 6.77 (6.08, 7.53) | 88.92 (84.95, 91.93) |
| Other prostheses | 3.10 (2.63, 3.65) | | | 0.48 (0.32, 0.71) | 3.57 (3.07, 4.16) | |
| Lower limb prostheses/orthoses | 2.23 (1.87, 2.66) | | | 0.12 (0.05, 0.27) | 2.35 (1.98, 2.79) | |
| Upper limb prosthetics/orthoses | 0.76 (0.54, 1.06) | | | 0.02 (0.01, 0.05) | 0.77 (0.55, 1.08) | |
| **Orthoses** | 7.04 (6.33, 7.82) | 6.98 (6.27, 7.76) | 0.06 (0.03, 0.11) | 0.59 (0.41, 0.87) | 7.63 (6.89, 8.44) | 91.43 (88.08, 93.90) |
| Other orthoses | 1.70 (1.37, 2.09) | | | 0.53 (0.35, 0.81) | 2.23 (1.85, 2.69) | |
| Lower limb orthoses | 4.80 (4.21, 5.47) | | | 0.05 (0.03, 0.10) | 4.85 (4.26, 5.52) | |
| Upper limb orthoses | 0.20 (0.11, 0.36) | | | 0.01 (0.00, 0.03) | 0.21 (0.12, 0.37) | |
| Spinal orthoses | 0.86 (0.66, 1.14) | | | 0.04 (0.02, 0.09) | 0.90 (0.69, 1.18) | |
| **AP for personal care** | 3.49 (3.15, 3.87) | 3.47 (3.13, 3.85) | 0.02 (0.01, 0.04) | 0.10 (0.06, 0.16) | 3.59 (3.24, 3.97) | 96.88 (95.07, 98.03) |
| AP to manage tissue integrity | 0.57 (0.46, 0.71) | | | 0.02 (0.01, 0.06) | 0.60 (0.48, 0.74) | |
| Absorbing products | 2.87 (2.56, 3.22) | | | 0.03 (0.01, 0.06) | 2.90 (2.59, 3.25) | |
| Catheter/device for collecting urine | 0.34 (0.25, 0.46) | | | 0.00 (0.00, 0.03) | 0.35 (0.26, 0.47) | |
| AP for ostomy care | 0.18 (0.11, 0.29) | | | 0.01 (0.00, 0.04) | 0.18 (0.11, 0.30) | |
| Adapted clothes, shoes | 0.38 (0.29, 0.50) | | | 0.05 (0.02, 0.11) | 0.43 (0.34, 0.56) | |
| **AP for everyday life** | 15.87 (14.86, 16.93) | 15.42 (14.43, 16.47) | 0.45 (0.32, 0.62) | 1.06 (0.80, 1.39) | 16.92 (15.89, 18.01) | 91.13 (89.09, 92.82) |
| Other products for everyday life | 1.08 (0.84, 1.38) | | | 0.37 (0.24, 0.59) | 1.45 (1.16, 1.81) | |
| Dentures | 13.02 (12.08, 14.02) | | | 0.56 (0.39, 0.82) | 13.58 (12.62, 14.60) | |
| AP for dressing | 0.80 (0.64, 1.02) | | | 0.08 (0.05, 0.15) | 0.89 (0.71, 1.11) | |
| AP for washing, bathing, showering | 1.91 (1.65, 2.20) | | | 0.22 (0.14, 0.36) | 2.13 (1.85, 2.44) | |
| AP for getting in and out of bed | 1.33 (1.16, 1.51) | | | 0.04 (0.02, 0.08) | 1.37 (1.20, 1.56) | |

| | Use | Met Need | Undermet Need | Unmet Need | Total Need | Coverage |
|---|---|---|---|---|---|---|
| AP for eating, drinking | 0.25 (0.17, 0.37) | | | 0.02 (0.01, 0.06) | 0.27 (0.18, 0.39) | |
| AP for domestic activities | 0.82 (0.67, 0.99) | | | 0.23 (0.13, 0.43) | 1.05 (0.86, 1.28) | |
| Personal emergency alarm system | 2.31 (2.02, 2.64) | | | 0.18 (0.10, 0.30) | 2.49 (2.18, 2.84) | |
| AP for toileting | 1.49 (1.26, 1.75) | | | 0.06 (0.04, 0.11) | 1.55 (1.33, 1.82) | |
| **AP for personal mobility** | 9.23 (8.59, 9.92) | 9.04 (8.40, 9.72) | 0.19 (0.14, 0.26) | 0.52 (0.35, 0.76) | 9.75 (9.08, 10.47) | 92.70 (90.37, 94.51) |
| Help from an animal | 0.01 (0.00, 0.03) | | | NA | 0.01 (0.00, 0.03) | |
| Other products for personal mobility | 0.22 (0.13, 0.38) | | | 0.05 (0.03, 0.08) | 0.27 (0.17, 0.42) | |
| White cane | 0.12 (0.08, 0.19) | | | 0.02 (0.01, 0.04) | 0.14 (0.10, 0.21) | |
| Canes, walking sticks | 7.56 (6.96, 8.21) | | | 0.44 (0.28, 0.69) | 8.01 (7.38, 8.68) | |
| Walker | 2.52 (2.26, 2.81) | | | 0.11 (0.07, 0.16) | 2.63 (2.36, 2.92) | |
| Electric wheelchair | 0.18 (0.14, 0.24) | | | 0.05 (0.03, 0.09) | 0.23 (0.18, 0.30) | |
| Manual wheelchair | 1.22 (1.07, 1.39) | | | 0.05 (0.03, 0.08) | 1.27 (1.12, 1.45) | |
| Tricycle, adapted bicycle, adapted scooter | 0.05 (0.03, 0.10) | | | 0.00 (0.00, 0.02) | 0.06 (0.03, 0.11) | |
| **AP for transfers, getting up, and going to bed** | 0.59 (0.47, 0.74) | 0.56 (0.44, 0.71) | 0.03 (0.01, 0.07) | 0.21 (0.16, 0.28) | 0.80 (0.66, 0.96) | 70.14 (61.92, 77.24) |
| Other products for changing position | 0.17 (0.11, 0.24) | | | 0.12 (0.08, 0.18) | 0.29 (0.22, 0.38) | |
| Patient lift, not fixed to the ceiling/walls | 0.30 (0.21, 0.43) | | | 0.03 (0.01, 0.05) | 0.32 (0.23, 0.46) | |
| Device to facilitate standing transfer | 0.12 (0.08, 0.17) | | | 0.05 (0.03, 0.09) | 0.16 (0.12, 0.23) | |
| Transfer board, lifting strap | 0.06 (0.03, 0.10) | | | 0.02 (0.00, 0.05) | 0.08 (0.05, 0.12) | |
| **AP for communication and to manage everyday life** | 0.79 (0.58, 1.08) | 0.78 (0.57, 1.06) | 0.01 (0.00, 0.04) | 0.28 (0.17, 0.44) | 1.07 (0.83, 1.38) | 72.92 (60.92, 82.30) |
| Device also used for other things | 0.10 (0.04, 0.27) | | | 0.07 (0.02, 0.24) | 0.16 (0.07, 0.36) | |
| Connected speakers | 0.01 (0.00, 0.04) | | | 0.01 (0.00, 0.03) | 0.02 (0.01, 0.05) | |

| | Use | Met Need | Undermet Need | Unmet Need | Total Need | Coverage |
|---|---|---|---|---|---|---|
| Input software/application | 0.01 (0.00, 0.03) | | | 0.00 (0.00, 0.02) | 0.01 (0.01, 0.03) | |
| Connected object | 0.08 (0.04, 0.15) | | | 0.01 (0.00, 0.04) | 0.09 (0.05, 0.16) | |
| Accessible input devices for computers/tablets | 0.22 (0.11, 0.43) | | | 0.12 (0.06, 0.25) | 0.34 (0.21, 0.56) | |
| Classic/adapted remote controls to open home/manage lights | 0.14 (0.06, 0.32) | | | 0.04 (0.02, 0.09) | 0.18 (0.09, 0.36) | |
| Adapted fixed network telephone | 0.21 (0.12, 0.35) | | | 0.07 (0.04, 0.12) | 0.28 (0.18, 0.42) | |
| Adapted mobile network telephone | 0.25 (0.15, 0.42) | | | 0.07 (0.04, 0.11) | 0.31 (0.21, 0.48) | |
| **Home Modifications** | 21.89 (20.71, 23.11) | 17.02 (15.94, 18.15) | 4.87 (4.38, 5.42) | 3.57 (3.07, 4.14) | 25.46 (24.19, 26.76) | 66.84 (64.30, 69.29) |
| **Toilet Area** | 10.15 (9.42, 10.94) | 9.62 (8.90, 10.39) | 0.54 (0.41, 0.69) | 1.91 (1.59, 2.28) | 12.06 (11.26, 12.91) | 79.75 (76.86, 82.37) |
| Supporting handrails, grab bars | 7.50 (6.89, 8.16) | | | 1.63 (1.37, 1.95) | 9.13 (8.46, 9.85) | |
| Area with enough space | 0.58 (0.43, 0.78) | | | 0.18 (0.13, 0.25) | 0.76 (0.60, 0.97) | |
| Single area for toilet, bathing areas | 0.65 (0.51, 0.84) | | | 0.21 (0.12, 0.36) | 0.86 (0.68, 1.09) | |
| Accessible electrical outlet/switch | 0.27 (0.17, 0.43) | | | 0.10 (0.06, 0.15) | 0.37 (0.26, 0.53) | |
| Floors with adapted materials | 0.70 (0.51, 0.96) | | | 0.31 (0.24, 0.41) | 1.01 (0.80, 1.28) | |
| Raised toilet seats/toilet seats with built-in raising mechanism | 4.35 (3.88, 4.88) | | | 0.99 (0.77, 1.27) | 5.34 (4.81, 5.92) | |
| **Bathing Area** | 16.12 (15.12, 17.16) | 14.39 (13.44, 15.39) | 1.73 (1.45, 2.06) | 4.27 (3.74, 4.88) | 20.39 (19.27, 21.55) | 70.57 (67.80, 73.19) |
| Supporting handrails, grab bars | 9.55 (8.80, 10.36) | | | 1.77 (1.48, 2.13) | 11.32 (10.51, 12.19) | |
| Enlarged shower unit/open shower stall | 9.15 (8.42, 9.94) | | | 4.38 (3.85, 4.99) | 13.54 (12.63, 14.49) | |
| Accessible sink | 0.41 (0.28, 0.59) | | | 0.24 (0.17, 0.33) | 0.65 (0.50, 0.84) | |
| Area with enough space | 1.03 (0.82, 1.29) | | | 0.36 (0.27, 0.50) | 1.39 (1.16, 1.67) | |
| Single area for toilet, bathing areas | 0.56 (0.39, 0.82) | | | 0.10 (0.07, 0.15) | 0.66 (0.48, 0.92) | |
| Accessible electrical outlet/switch | 0.69 (0.49, 0.96) | | | 0.16 (0.11, 0.23) | 0.84 (0.63, 1.12) | |

| | Use | Met Need | Undermet Need | Unmet Need | Total Need | Coverage |
|---|---|---|---|---|---|---|
| Bath/shower seat fixed to the wall | 4.21 (3.76, 4.71) | | | 1.51 (1.25, 1.83) | 5.72 (5.19, 6.30) | |
| Floors with adapted materials | 1.91 (1.58, 2.30) | | | 0.59 (0.43, 0.81) | 2.50 (2.13, 2.94) | |
| **Kitchen** | 1.05 (0.81, 1.35) | 1.00 (0.77, 1.30) | 0.04 (0.02, 0.09) | 0.64 (0.49, 0.83) | 1.68 (1.40, 2.03) | 59.47 (50.46, 67.89) |
| Specialized equipment | 0.05 (0.03, 0.09) | | | 0.09 (0.05, 0.13) | 0.14 (0.10, 0.19) | |
| Accessible sink, lowered/height-adjustable | 0.14 (0.07, 0.26) | | | 0.19 (0.12, 0.31) | 0.33 (0.22, 0.48) | |
| Specialized furniture | 0.15 (0.09, 0.25) | | | 0.15 (0.11, 0.22) | 0.30 (0.22, 0.42) | |
| Counters, lowered/height-adjustable | 0.33 (0.20, 0.54) | | | 0.22 (0.13, 0.37) | 0.55 (0.39, 0.79) | |
| Accessible electrical outlet/switch | 0.42 (0.28, 0.64) | | | 0.09 (0.06, 0.14) | 0.51 (0.36, 0.73) | |
| Ramp/handrail/grab bar | 0.14 (0.09, 0.21) | | | 0.19 (0.13, 0.28) | 0.32 (0.24, 0.43) | |
| Floors with adapted materials | 0.02 (0.01, 0.05) | | | 0.12 (0.08, 0.19) | 0.15 (0.10, 0.22) | |
| **Stairs** | 7.93 (7.17, 8.76) | 7.50 (6.76, 8.32) | 0.42 (0.30, 0.60) | 1.27 (1.04, 1.56) | 9.20 (8.41, 10.07) | 81.53 (78.23, 84.44) |
| Elevator | 1.17 (0.83, 1.65) | | | 0.31 (0.21, 0.45) | 1.48 (1.12, 1.96) | |
| Stairlifts with seat | 0.83 (0.61, 1.12) | | | 0.98 (0.77, 1.24) | 1.80 (1.49, 2.18) | |
| Platform lift | 0.05 (0.02, 0.16) | | | 0.08 (0.04, 0.18) | 0.14 (0.07, 0.26) | |
| One/two supporting handrails | 6.42 (5.75, 7.15) | | | 0.48 (0.35, 0.65) | 6.89 (6.21, 7.64) | |
| **Other areas of the residence** | 4.24 (3.74, 4.80) | 4.03 (3.54, 4.58) | 0.22 (0.13, 0.35) | 1.06 (0.86, 1.31) | 5.31 (4.76, 5.91) | 75.87 (71.38, 79.86) |
| Supporting handrails, grab bars | 1.13 (0.88, 1.46) | | | 0.37 (0.27, 0.50) | 1.50 (1.22, 1.84) | |
| Light device to alert to noises | 0.05 (0.03, 0.11) | | | 0.02 (0.01, 0.06) | 0.08 (0.04, 0.13) | |
| Motorized windows/electric curtains | 2.31 (1.96, 2.72) | | | 0.62 (0.46, 0.84) | 2.93 (2.54, 3.38) | |
| Stationary hoists fixed to ceiling/walls | 0.11 (0.05, 0.26) | | | 0.03 (0.02, 0.06) | 0.15 (0.08, 0.28) | |
| Automatic light fixtures/light strip | 0.50 (0.35, 0.71) | | | 0.20 (0.14, 0.28) | 0.70 (0.53, 0.92) | |

| | Use | Met Need | Undermet Need | Unmet Need | Total Need | Coverage |
|---|---|---|---|---|---|---|
| Other furniture, lowered/height-adjustable | 0.11 (0.06, 0.18) | | | 0.09 (0.05, 0.14) | 0.19 (0.14, 0.28) | |
| Automatic doors | 0.21 (0.11, 0.41) | | | 0.06 (0.03, 0.10) | 0.27 (0.16, 0.46) | |
| Enlarged doors/hallways | 0.48 (0.35, 0.66) | | | 0.16 (0.08, 0.29) | 0.64 (0.48, 0.85) | |
| Non-slip floor coverings | 0.17 (0.10, 0.30) | | | 0.11 (0.07, 0.16) | 0.28 (0.19, 0.41) | |
| **Human Help** | 18.53 (17.56, 19.54) | 13.45 (12.62, 14.33) | 5.08 (4.59, 5.62) | 3.08 (2.61, 3.63) | 21.61 (20.54, 22.72) | 62.23 (59.61, 64.78) |
| Having regular, balanced diet | 2.05 (1.81, 2.32) | 1.99 (1.75, 2.25) | 0.07 (0.04, 0.11) | 0.25 (0.15, 0.39) | 2.30 (2.04, 2.59) | 86.45 (80.96, 90.54) |
| Managing money, budget | 3.13 (2.81, 3.49) | 3.07 (2.75, 3.42) | 0.06 (0.04, 0.11) | 0.14 (0.09, 0.21) | 3.27 (2.94, 3.63) | 93.89 (91.52, 95.63) |
| Finding one's path | 1.40 (1.20, 1.63) | 1.37 (1.17, 1.61) | 0.03 (0.01, 0.05) | 0.12 (0.05, 0.28) | 1.52 (1.30, 1.78) | 90.40 (81.97, 95.12) |
| Cutting food/serving drinks | 1.80 (1.59, 2.05) | 1.73 (1.52, 1.97) | 0.07 (0.04, 0.12) | 0.09 (0.05, 0.14) | 1.89 (1.67, 2.14) | 91.60 (88.24, 94.06) |
| Doing groceries | 10.12 (9.50, 10.76) | 9.31 (8.72, 9.94) | 0.80 (0.65, 0.99) | 0.90 (0.70, 1.15) | 11.01 (10.36, 11.70) | 84.56 (82.11, 86.73) |
| Arranging day/night care | 1.46 (1.27, 1.68) | 1.38 (1.19, 1.60) | 0.08 (0.05, 0.12) | 0.16 (0.11, 0.23) | 1.61 (1.41, 1.84) | 85.55 (80.82, 89.27) |
| Getting dressed/undressed | 3.28 (3.00, 3.59) | 2.95 (2.70, 3.23) | 0.33 (0.22, 0.49) | 0.16 (0.10, 0.27) | 3.44 (3.15, 3.76) | 85.75 (81.26, 89.31) |
| Bathing | 3.83 (3.51, 4.19) | 3.40 (3.10, 3.71) | 0.44 (0.31, 0.62) | 0.29 (0.18, 0.45) | 4.12 (3.77, 4.50) | 82.44 (77.77, 86.30) |
| Lying down in bed/rising from bed | 1.46 (1.28, 1.66) | 1.29 (1.14, 1.45) | 0.17 (0.09, 0.32) | 0.06 (0.04, 0.11) | 1.52 (1.34, 1.72) | 84.74 (77.08, 90.16) |
| Eating/drinking, once food has been served or cut | 0.49 (0.40, 0.60) | 0.47 (0.38, 0.58) | 0.02 (0.01, 0.05) | 0.04 (0.02, 0.08) | 0.52 (0.43, 0.64) | 89.72 (81.93, 94.38) |
| Taking medication | 2.78 (2.51, 3.08) | 2.71 (2.44, 3.00) | 0.08 (0.05, 0.13) | 0.13 (0.06, 0.29) | 2.91 (2.62, 3.23) | 92.97 (88.32, 95.86) |
| Using mode of transportation | 5.76 (5.30, 6.26) | 5.36 (4.92, 5.83) | 0.40 (0.29, 0.55) | 0.47 (0.34, 0.66) | 6.23 (5.75, 6.75) | 85.96 (82.70, 88.69) |
| Using computer/tablet/other electronic device | 1.98 (1.64, 2.39) | 1.83 (1.50, 2.22) | 0.15 (0.08, 0.29) | 1.35 (1.02, 1.77) | 3.33 (2.84, 3.89) | 54.89 (46.78, 62.76) |
| Managing medical appointments | 4.55 (4.14, 5.00) | 4.36 (3.96, 4.79) | 0.19 (0.11, 0.34) | 0.24 (0.17, 0.35) | 4.79 (4.37, 5.25) | 90.88 (87.73, 93.28) |
| Preparing meals | 4.25 (3.89, 4.64) | 4.05 (3.69, 4.43) | 0.20 (0.15, 0.27) | 0.40 (0.27, 0.59) | 4.65 (4.26, 5.07) | 87.07 (83.42, 90.01) |
| Moving around all areas of floor | 0.72 (0.60, 0.85) | 0.69 (0.58, 0.82) | 0.03 (0.01, 0.06) | 0.07 (0.02, 0.20) | 0.78 (0.65, 0.94) | 87.89 (75.97, 94.34) |

| | Use | Met Need | Undermet Need | Unmet Need | Total Need | Coverage |
|---|---|---|---|---|---|---|
| Sitting down/standing up from chair | 1.07 (0.92, 1.24) | 0.97 (0.84, 1.11) | 0.10 (0.04, 0.25) | 0.07 (0.02, 0.18) | 1.13 (0.97, 1.32) | 85.19 (74.21, 92.01) |
| Leaving home | 3.45 (3.14, 3.78) | 3.25 (2.95, 3.58) | 0.19 (0.14, 0.26) | 0.21 (0.15, 0.29) | 3.66 (3.34, 4.00) | 88.95 (86.41, 91.06) |
| Executing usual administrative procedures | 7.67 (7.10, 8.29) | 7.22 (6.66, 7.82) | 0.46 (0.34, 0.62) | 1.21 (0.92, 1.59) | 8.89 (8.23, 9.59) | 81.21 (77.43, 84.48) |
| Running everyday errands/tasks in the home | 9.59 (8.98, 10.24) | 8.11 (7.54, 8.71) | 1.48 (1.27, 1.73) | 2.33 (1.99, 2.73) | 11.92 (11.21, 12.67) | 68.01 (65.00, 70.88) |
| Running less frequent tasks | 10.33 (9.66, 11.04) | 9.07 (8.45, 9.74) | 1.26 (1.04, 1.52) | 2.01 (1.63, 2.47) | 12.34 (11.56, 13.16) | 73.53 (70.20, 76.62) |
| Using telephone | 1.28 (1.09, 1.51) | 1.26 (1.07, 1.49) | 0.02 (0.01, 0.06) | 0.20 (0.10, 0.41) | 1.49 (1.25, 1.76) | 84.78 (73.97, 91.61) |
| Using the toilet | 0.81 (0.70, 0.94) | 0.78 (0.67, 0.91) | 0.03 (0.02, 0.07) | 0.03 (0.02, 0.07) | 0.85 (0.73, 0.98) | 92.24 (87.41, 95.32) |
| Choosing appropriate clothes | 1.24 (1.05, 1.45) | 1.20 (1.02, 1.42) | 0.03 (0.01, 0.07) | 0.12 (0.05, 0.24) | 1.35 (1.15, 1.59) | 89.13 (81.31, 93.92) |

Use: the proportion of the population reporting use of a particular tool – an assistive product, home modification or human help**,** Met Need: the proportion of the population whose need is fully satisfied by the tool they use, Undermet Need: the proportion of the population that use a tool but report needing an updated or different tool within the same group category, Unmet Need: the proportion of the population who reports needing a tool but does not use one, Total Need: the proportion of the population that either use or report needing the tool, Coverage: the proportion of individuals who use or need a tool whose need is met

**Table S11: Reasons for not having an assistive product that is needed in participants aged 60 or older.**

| | Weighted percentage (Observed Frequency) |
|---|---|
| **Does not have the assistive products they need,** | 7.7 (914) |
| **Reasons** | |
| It is too expensive | 35.4 (334) |
| It is hard to find, obtain (availability, administrative hurdles, lack of information) | 9.8 (104) |
| It is too constraining, difficult to use | 8.4 (82) |
| These products are for people whose health is in a worse state than yours | 6.4 (59) |
| You hadn’t thought about it | 6.9 (76) |
| You prefer not using one | 15.6 (151) |
| Using one made your health problems too obvious | 1.5 (25) |
| You don’t believe this will help in the long-run | 4.7 (38) |
| Other reason | 22.8 (218) |

Statistics are weighted percentage for target population (observed frequency among survey respondents) unless specified otherwise.

The weighted percentages under “Reasons” express the estimated proportion of people selecting each reason among those who declared needing an assistive product they did not have. Participants were able to select multiple reasons in this list.

**Table S12: Reasons for not having a home modification that is needed in participants aged 60 or older.**

| | Weighted percentage (Observed Frequency) |
|---|---|
| **Does not have a home modification they need** | 8.4 (1,335) |
| **Reasons** | |
| It is too expensive | 40.3 (605) |
| It is hard to obtain (availability, administrative hurdles, lack of information) | 14.6 (240) |
| It is too constraining, difficult to use | 1.5 (16) |
| These products are for people whose health is in a worse state than yours | 8.1 (80) |
| You hadn't thought about it | 12.5 (155) |
| You are scared of damaging your home | 2.7 (43) |
| The people you live with don't want these | 1.0 (24) |
| It is not possible to make these modifications in your current home | 14.9 (190) |
| You don't believe this will help in the long-run | 1.6 (21) |

Statistics are weighted percentage for target population (observed frequency among survey respondents) unless specified otherwise.

The weighted percentages under "Reasons" express the estimated proportion of people selecting each reason among those who declared needing a home modification they did not have. Participants were able to select multiple reasons in this list.

**Table S13. Use for General Population and Socio-demographic Subgroups [Socio-demographic]**

| | Total population | Socio-demographic Subgroups | | | |
|---|---|---|---|---|---|
| | | Women | Low-education | Oldest-old | Renounced to care |
| **AP** | 80.62 (79.14, 82.01) | 81.49 (79.47, 83.36) | 81.71 (79.98, 83.33) | 90.60 (88.42, 92.41) | 87.99 (83.80, 91.21) |
| **AP for seeing, reading** | 75.52 (74.00, 76.99) | 76.05 (73.96, 78.02) | 75.85 (74.04, 77.58) | 79.20 (76.46, 81.69) | 81.55 (77.19, 85.25) |
| **AP for hearing, speaking** | 11.59 (10.64, 12.60) | 9.71 (8.59, 10.97) | 12.54 (11.38, 13.81) | 26.93 (24.12, 29.93) | 10.96 (8.40, 14.18) |
| **Prosthetics and implants** | 6.08 (5.43, 6.81) | 6.54 (5.63, 7.58) | 6.64 (5.84, 7.54) | 9.41 (7.88, 11.19) | 6.89 (5.16, 9.14) |
| **Orthoses** | 7.04 (6.33, 7.82) | 8.25 (7.22, 9.40) | 7.13 (6.30, 8.07) | 9.03 (7.62, 10.66) | 11.33 (8.83, 14.44) |
| **AP for personal care** | 3.49 (3.15, 3.87) | 4.41 (3.89, 4.99) | 4.39 (3.93, 4.91) | 11.73 (10.40, 13.21) | 6.32 (4.84, 8.22) |
| **AP for everyday life** | 15.87 (14.86, 16.93) | 17.15 (15.76, 18.64) | 19.90 (18.54, 21.33) | 36.06 (33.23, 38.99) | 19.02 (15.99, 22.47) |
| **AP for personal mobility** | 9.23 (8.59, 9.92) | 10.90 (9.95, 11.92) | 11.19 (10.33, 12.10) | 27.60 (25.20, 30.14) | 17.59 (14.56, 21.10) |
| **AP for transfers, getting up, and going to bed** | 0.59 (0.47, 0.74) | 0.63 (0.48, 0.82) | 0.75 (0.57, 0.98) | 2.15 (1.59, 2.92) | 1.09 (0.68, 1.73) |
| **AP for communication and to manage everyday life** | 0.79 (0.58, 1.08) | 0.95 (0.65, 1.39) | 1.00 (0.69, 1.43) | 2.49 (1.67, 3.70) | 0.83 (0.51, 1.35) |
| **Home Modifications** | 21.89 (20.71, 23.11) | 24.93 (23.23, 26.71) | 25.02 (23.54, 26.55) | 50.11 (46.88, 53.33) | 27.63 (23.94, 31.65) |
| **Toilet Area** | 10.15 (9.42, 10.94) | 11.83 (10.77, 13.00) | 12.59 (11.59, 13.66) | 28.28 (25.80, 30.89) | 14.15 (11.76, 16.94) |
| **Bathing Area** | 16.12 (15.12, 17.16) | 18.46 (17.03, 19.99) | 18.87 (17.58, 20.22) | 38.11 (35.19, 41.11) | 19.42 (16.54, 22.67) |
| **Kitchen** | 1.05 (0.81, 1.35) | 1.28 (0.92, 1.76) | 1.04 (0.76, 1.40) | 2.14 (1.39, 3.28) | 1.11 (0.74, 1.66) |
| **Stairs** | 7.93 (7.17, 8.76) | 8.56 (7.50, 9.76) | 9.08 (8.13, 10.12) | 18.23 (16.00, 20.70) | 9.42 (7.08, 12.44) |
| **Other areas of the residence** | 4.24 (3.74, 4.80) | 4.88 (4.17, 5.71) | 4.66 (4.04, 5.38) | 9.96 (8.40, 11.79) | 5.54 (4.12, 7.40) |
| **Human Help** | 18.53 (17.56, 19.54) | 23.01 (21.52, 24.57) | 22.85 (21.54, 24.21) | 52.21 (48.94, 55.46) | 27.83 (24.28, 31.69) |
| Having regular, balanced diet | 2.05 (1.81, 2.32) | 2.24 (1.93, 2.60) | 2.62 (2.27, 3.01) | 6.87 (5.87, 8.02) | 3.73 (2.63, 5.27) |
| Managing money, budget | 3.13 (2.81, 3.49) | 3.65 (3.18, 4.17) | 4.14 (3.68, 4.65) | 10.82 (9.52, 12.29) | 5.05 (3.79, 6.69) |

| | Total population | Socio-demographic Subgroups | | | |
|---|---|---|---|---|---|
| | | Women | Low-education | Oldest-old | Renounced to care |
| Finding one's path | 1.40 (1.20, 1.63) | 1.77 (1.46, 2.14) | 1.71 (1.45, 2.01) | 3.92 (3.29, 4.67) | 2.08 (1.50, 2.89) |
| Cutting food/serving drinks | 1.80 (1.59, 2.05) | 2.16 (1.84, 2.53) | 2.33 (2.02, 2.68) | 5.77 (4.90, 6.79) | 3.11 (2.37, 4.07) |
| Doing groceries | 10.12 (9.50, 10.76) | 13.16 (12.21, 14.18) | 13.03 (12.15, 13.96) | 31.13 (28.64, 33.73) | 16.24 (13.91, 18.88) |
| Arranging day/night care | 1.46 (1.27, 1.68) | 1.59 (1.34, 1.90) | 1.89 (1.61, 2.22) | 4.60 (3.93, 5.36) | 2.39 (1.77, 3.22) |
| Getting dressed/undressed | 3.28 (3.00, 3.59) | 3.71 (3.31, 4.16) | 4.23 (3.83, 4.67) | 10.06 (8.94, 11.31) | 5.83 (4.75, 7.15) |
| Bathing | 3.83 (3.51, 4.19) | 4.43 (3.98, 4.93) | 4.99 (4.54, 5.48) | 12.76 (11.38, 14.29) | 6.32 (5.19, 7.68) |
| Lying down in bed/rising from bed | 1.46 (1.28, 1.66) | 1.67 (1.43, 1.96) | 1.89 (1.63, 2.18) | 5.14 (4.32, 6.11) | 3.04 (2.29, 4.02) |
| Eating/drinking, once food has been served or cut | 0.49 (0.40, 0.60) | 0.59 (0.46, 0.77) | 0.65 (0.52, 0.81) | 1.72 (1.32, 2.24) | 0.94 (0.55, 1.60) |
| Taking medication | 2.78 (2.51, 3.08) | 3.13 (2.74, 3.58) | 3.57 (3.19, 3.99) | 9.34 (8.26, 10.54) | 4.18 (3.26, 5.34) |
| Using mode of transportation | 5.76 (5.30, 6.26) | 7.36 (6.67, 8.12) | 7.35 (6.71, 8.03) | 17.99 (16.19, 19.94) | 10.04 (8.01, 12.53) |
| Using computer/tablet/other electronic device | 1.98 (1.64, 2.39) | 2.33 (1.82, 2.97) | 2.46 (1.99, 3.05) | 5.61 (4.31, 7.28) | 2.37 (1.46, 3.83) |
| Managing medical appointments | 4.55 (4.14, 5.00) | 5.00 (4.45, 5.62) | 6.06 (5.47, 6.71) | 16.38 (14.61, 18.31) | 7.10 (5.48, 9.15) |
| Preparing meals | 4.25 (3.89, 4.64) | 5.00 (4.50, 5.56) | 5.43 (4.91, 6.00) | 13.56 (12.05, 15.23) | 7.00 (5.57, 8.77) |
| Moving around all areas of floor | 0.72 (0.60, 0.85) | 0.87 (0.70, 1.08) | 0.90 (0.74, 1.08) | 2.24 (1.81, 2.78) | 1.37 (0.92, 2.04) |
| Sitting down/standing up from chair | 1.07 (0.92, 1.24) | 1.21 (1.01, 1.44) | 1.35 (1.14, 1.61) | 3.50 (2.84, 4.32) | 2.21 (1.60, 3.06) |
| Leaving home | 3.45 (3.14, 3.78) | 4.62 (4.17, 5.11) | 4.43 (4.01, 4.90) | 11.01 (9.78, 12.37) | 6.25 (4.79, 8.10) |
| Executing usual administrative procedures | 7.67 (7.10, 8.29) | 9.28 (8.42, 10.21) | 10.26 (9.43, 11.15) | 27.23 (24.77, 29.83) | 10.30 (8.42, 12.55) |
| Running everyday errands/tasks in the home | 9.59 (8.98, 10.24) | 12.46 (11.53, 13.47) | 11.85 (11.01, 12.74) | 28.51 (26.16, 30.99) | 15.79 (13.42, 18.48) |
| Running less frequent tasks | 10.33 (9.66, 11.04) | 13.82 (12.75, 14.98) | 12.77 (11.86, 13.74) | 30.38 (27.87, 33.01) | 15.78 (13.44, 18.44) |
| Using telephone | 1.28 (1.09, 1.51) | 1.33 (1.08, 1.64) | 1.66 (1.39, 1.98) | 3.75 (3.10, 4.54) | 2.12 (1.34, 3.34) |

| | Total population | Socio-demographic Subgroups | | | |
|---|---|---|---|---|---|
| | | Women | Low-education | Oldest-old | Renounced to care |
| Using the toilet | 0.81 (0.70, 0.94) | 0.98 (0.81, 1.19) | 1.06 (0.90, 1.25) | 2.75 (2.26, 3.34) | 1.94 (1.42, 2.66) |
| Choosing appropriate clothes | 1.24 (1.05, 1.45) | 1.17 (0.94, 1.45) | 1.49 (1.25, 1.77) | 3.91 (3.20, 4.77) | 2.51 (1.55, 4.02) |

**Table S14. Use for General Population and Morbidity Subgroups**

| | General | Morbidity Subgroups | | |
|---|---|---|---|---|
| | | Frail | Any Chronic Condition | Multimorbid |
| **AP** | 80.62 (79.14, 82.01) | 88.99 (87.12, 90.62) | 84.49 (83.03, 85.85) | 87.27 (85.71, 88.69) |
| **AP for seeing, reading** | 75.52 (74.00, 76.99) | 77.17 (74.98, 79.22) | 78.48 (76.92, 79.96) | 80.83 (79.13, 82.43) |
| **AP for hearing, speaking** | 11.59 (10.64, 12.60) | 18.34 (16.49, 20.35) | 13.38 (12.27, 14.58) | 15.41 (14.05, 16.86) |
| **Prosthetics and implants** | 6.08 (5.43, 6.81) | 10.18 (8.92, 11.59) | 7.16 (6.38, 8.03) | 8.43 (7.46, 9.52) |
| **Orthoses** | 7.04 (6.33, 7.82) | 12.80 (11.41, 14.33) | 8.50 (7.65, 9.44) | 10.56 (9.47, 11.76) |
| **AP for personal care** | 3.49 (3.15, 3.87) | 11.72 (10.60, 12.93) | 4.31 (3.88, 4.78) | 5.60 (5.02, 6.25) |
| **AP for everyday life** | 15.87 (14.86, 16.93) | 32.54 (30.50, 34.66) | 18.31 (17.14, 19.54) | 21.91 (20.45, 23.44) |
| **AP for personal mobility** | 9.23 (8.59, 9.92) | 32.21 (30.15, 34.34) | 11.39 (10.58, 12.24) | 14.46 (13.41, 15.58) |
| **AP for transfers, getting up, and going to bed** | 0.59 (0.47, 0.74) | 2.31 (1.82, 2.91) | 0.73 (0.57, 0.92) | 0.94 (0.73, 1.21) |
| **AP for communication and to manage everyday life** | 0.79 (0.58, 1.08) | 1.85 (1.41, 2.41) | 0.94 (0.69, 1.27) | 1.16 (0.85, 1.59) |
| **Home Modifications** | 21.89 (20.71, 23.11) | 48.91 (46.48, 51.34) | 26.15 (24.76, 27.60) | 31.29 (29.57, 33.06) |
| **Toilet Area** | 10.15 (9.42, 10.94) | 29.14 (27.15, 31.20) | 12.43 (11.52, 13.39) | 15.75 (14.56, 17.01) |
| **Bathing Area** | 16.12 (15.12, 17.16) | 38.27 (36.02, 40.57) | 19.41 (18.21, 20.67) | 23.74 (22.22, 25.32) |
| **Kitchen** | 1.05 (0.81, 1.35) | 2.41 (1.80, 3.23) | 1.23 (0.95, 1.59) | 1.46 (1.13, 1.90) |
| **Stairs** | 7.93 (7.17, 8.76) | 16.51 (14.90, 18.26) | 9.33 (8.44, 10.32) | 11.04 (9.95, 12.24) |
| **Other areas of the residence** | 4.24 (3.74, 4.80) | 9.55 (8.41, 10.82) | 5.18 (4.57, 5.86) | 6.33 (5.57, 7.19) |
| **Human Help** | 18.53 (17.56, 19.54) | 53.78 (51.28, 56.26) | 22.47 (21.30, 23.69) | 27.66 (26.15, 29.22) |
| Having regular, balanced diet | 2.05 (1.81, 2.32) | 7.22 (6.38, 8.16) | 2.50 (2.21, 2.84) | 3.06 (2.69, 3.47) |
| Managing money, budget | 3.13 (2.81, 3.49) | 10.30 (9.25, 11.46) | 3.81 (3.41, 4.26) | 4.57 (4.07, 5.12) |

| | | | | |
|---|---|---|---|---|
| Finding one's path | 1.40 (1.20, 1.63) | 4.62 (3.94, 5.40) | 1.69 (1.44, 1.98) | 1.93 (1.64, 2.27) |
| Cutting food/serving drinks | 1.80 (1.59, 2.05) | 6.71 (5.93, 7.57) | 2.21 (1.94, 2.51) | 2.81 (2.45, 3.23) |
| Doing groceries | 10.12 (9.50, 10.76) | 35.21 (33.16, 37.32) | 12.39 (11.63, 13.20) | 15.57 (14.57, 16.63) |
| Arranging day/night care | 1.46 (1.27, 1.68) | 5.44 (4.71, 6.27) | 1.77 (1.53, 2.04) | 2.19 (1.88, 2.55) |
| Getting dressed/undressed | 3.28 (3.00, 3.59) | 12.46 (11.39, 13.62) | 4.03 (3.67, 4.41) | 5.19 (4.71, 5.73) |
| Bathing | 3.83 (3.51, 4.19) | 14.54 (13.33, 15.85) | 4.68 (4.27, 5.13) | 5.90 (5.35, 6.50) |
| Lying down in bed/rising from bed | 1.46 (1.28, 1.66) | 5.79 (5.07, 6.60) | 1.80 (1.58, 2.05) | 2.26 (1.96, 2.61) |
| Eating/drinking, once food has been served or cut | 0.49 (0.40, 0.60) | 1.93 (1.57, 2.37) | 0.60 (0.49, 0.74) | 0.68 (0.54, 0.86) |
| Taking medication | 2.78 (2.51, 3.08) | 9.55 (8.60, 10.59) | 3.42 (3.08, 3.79) | 4.11 (3.69, 4.58) |
| Using mode of transportation | 5.76 (5.30, 6.26) | 19.26 (17.75, 20.87) | 7.02 (6.46, 7.63) | 8.56 (7.85, 9.32) |
| Using computer/tablet/other electronic device | 1.98 (1.64, 2.39) | 5.32 (4.32, 6.55) | 2.36 (1.95, 2.85) | 2.65 (2.16, 3.23) |
| Managing medical appointments | 4.55 (4.14, 5.00) | 14.72 (13.43, 16.11) | 5.57 (5.06, 6.13) | 6.78 (6.14, 7.48) |
| Preparing meals | 4.25 (3.89, 4.64) | 15.42 (14.14, 16.81) | 5.21 (4.76, 5.70) | 6.49 (5.91, 7.12) |
| Moving around all areas of floor | 0.72 (0.60, 0.85) | 2.83 (2.38, 3.35) | 0.88 (0.74, 1.04) | 1.11 (0.92, 1.33) |
| Sitting down/standing up from chair | 1.07 (0.92, 1.24) | 4.25 (3.65, 4.95) | 1.31 (1.12, 1.52) | 1.66 (1.41, 1.96) |
| Leaving home | 3.45 (3.14, 3.78) | 13.42 (12.22, 14.71) | 4.22 (3.84, 4.64) | 5.46 (4.94, 6.03) |
| Executing usual administrative procedures | 7.67 (7.10, 8.29) | 23.23 (21.52, 25.03) | 9.26 (8.56, 10.02) | 11.15 (10.27, 12.08) |
| Running everyday errands/tasks in the home | 9.59 (8.98, 10.24) | 32.73 (30.77, 34.75) | 11.76 (11.00, 12.56) | 14.79 (13.79, 15.86) |
| Running less frequent tasks | 10.33 (9.66, 11.04) | 32.23 (30.27, 34.26) | 12.56 (11.74, 13.43) | 15.84 (14.76, 16.99) |
| Using telephone | 1.28 (1.09, 1.51) | 3.72 (3.21, 4.32) | 1.58 (1.34, 1.86) | 1.85 (1.56, 2.20) |
| Using the toilet | 0.81 (0.70, 0.94) | 3.25 (2.79, 3.78) | 0.99 (0.85, 1.15) | 1.22 (1.04, 1.44) |
| Choosing appropriate clothes | 1.24 (1.05, 1.45) | 4.03 (3.51, 4.63) | 1.51 (1.28, 1.78) | 1.65 (1.43, 1.90) |

**Table S15. Use for General Population and Limitations Subgroups**

| | General | Limitations Subgroups | | | | |
|---|---|---|---|---|---|---|
| | | GIR | ADL/IADL | ADL | IADL | Self-reported limitations |
| **AP** | 80.62 (79.14, 82.01) | 94.08 (92.29, 95.48) | 90.31 (88.62, 91.77) | 93.26 (91.80, 94.49) | 89.98 (88.15, 91.55) | 85.91 (84.17, 87.49) |
| **AP for seeing, reading** | 75.52 (74.00, 76.99) | 73.23 (69.87, 76.34) | 78.19 (76.04, 80.20) | 75.82 (73.07, 78.36) | 78.04 (75.72, 80.20) | 76.57 (74.60, 78.43) |
| **AP for hearing, speaking** | 11.59 (10.64, 12.60) | 19.69 (16.99, 22.71) | 17.39 (15.66, 19.27) | 19.27 (16.90, 21.88) | 17.78 (15.91, 19.82) | 15.50 (14.04, 17.08) |
| **Prosthetics and implants** | 6.08 (5.43, 6.81) | 11.52 (9.53, 13.86) | 10.71 (9.38, 12.21) | 12.20 (10.42, 14.23) | 10.61 (9.21, 12.20) | 9.37 (8.26, 10.60) |
| **Orthoses** | 7.04 (6.33, 7.82) | 17.89 (15.37, 20.71) | 13.42 (11.98, 15.02) | 16.49 (14.48, 18.72) | 13.36 (11.85, 15.02) | 11.94 (10.70, 13.31) |
| **AP for personal care** | 3.49 (3.15, 3.87) | 27.70 (25.02, 30.54) | 10.60 (9.58, 11.71) | 19.24 (17.34, 21.30) | 9.57 (8.55, 10.69) | 8.04 (7.25, 8.91) |
| **AP for everyday life** | 15.87 (14.86, 16.93) | 48.84 (45.47, 52.23) | 31.20 (29.18, 33.29) | 42.59 (39.78, 45.44) | 30.68 (28.55, 32.89) | 26.05 (24.35, 27.83) |
| **AP for personal mobility** | 9.23 (8.59, 9.92) | 53.05 (49.62, 56.45) | 28.60 (26.69, 30.59) | 42.99 (40.17, 45.86) | 27.93 (25.91, 30.04) | 21.39 (19.92, 22.93) |
| **AP for transfers, getting up, and going to bed** | 0.59 (0.47, 0.74) | 6.76 (5.28, 8.62) | 2.06 (1.62, 2.60) | 4.30 (3.39, 5.45) | 1.89 (1.45, 2.48) | 1.52 (1.21, 1.92) |
| **AP for communication and to manage everyday life** | 0.79 (0.58, 1.08) | 4.13 (2.94, 5.79) | 2.16 (1.60, 2.91) | 2.80 (2.05, 3.82) | 2.26 (1.65, 3.09) | 1.86 (1.36, 2.54) |
| **Home Modifications** | 21.89 (20.71, 23.11) | 63.72 (60.13, 67.17) | 46.48 (44.08, 48.89) | 59.25 (56.23, 62.20) | 45.64 (43.10, 48.20) | 38.11 (36.11, 40.16) |
| **Toilet Area** | 10.15 (9.42, 10.94) | 43.75 (40.52, 47.05) | 26.45 (24.59, 28.40) | 37.65 (35.00, 40.38) | 25.15 (23.22, 27.18) | 20.61 (19.14, 22.15) |
| **Bathing Area** | 16.12 (15.12, 17.16) | 50.02 (46.61, 53.42) | 36.16 (33.97, 38.40) | 45.18 (42.30, 48.08) | 35.37 (33.06, 37.74) | 28.96 (27.19, 30.80) |
| **Kitchen** | 1.05 (0.81, 1.35) | 3.84 (2.66, 5.52) | 2.35 (1.79, 3.08) | 3.98 (2.87, 5.50) | 2.27 (1.70, 3.03) | 2.09 (1.60, 2.74) |
| **Stairs** | 7.93 (7.17, 8.76) | 19.60 (17.23, 22.19) | 15.90 (14.29, 17.65) | 20.35 (18.07, 22.84) | 15.58 (13.91, 17.40) | 13.55 (12.20, 15.02) |
| **Other areas of the residence** | 4.24 (3.74, 4.80) | 17.02 (14.72, 19.59) | 10.23 (8.98, 11.63) | 13.70 (11.98, 15.61) | 10.11 (8.79, 11.61) | 7.90 (6.94, 8.98) |
| **Human Help** | 18.53 (17.56, 19.54) | 82.11 (78.63, 85.12) | 59.41 (56.87, 61.90) | 70.73 (67.57, 73.71) | 59.85 (57.14, 62.51) | 39.44 (37.48, 41.44) |
| Having regular, balanced diet | 2.05 (1.81, 2.32) | 19.16 (17.07, 21.45) | 7.09 (6.24, 8.03) | 13.07 (11.57, 14.72) | 6.06 (5.22, 7.03) | 4.88 (4.30, 5.52) |
| Managing money, budget | 3.13 (2.81, 3.49) | 25.85 (23.21, 28.69) | 10.73 (9.60, 11.96) | 17.88 (16.02, 19.90) | 9.85 (8.69, 11.15) | 7.28 (6.52, 8.13) |

| | | | | | | |
|---|---|---|---|---|---|---|
| Finding one's path | 1.40 (1.20, 1.63) | 10.95 (9.38, 12.74) | 4.90 (4.19, 5.73) | 7.50 (6.48, 8.67) | 4.62 (3.88, 5.50) | 3.17 (2.75, 3.66) |
| Cutting food/serving drinks | 1.80 (1.59, 2.05) | 21.93 (19.41, 24.67) | 6.35 (5.59, 7.21) | 13.09 (11.51, 14.85) | 5.55 (4.78, 6.44) | 4.44 (3.93, 5.02) |
| Doing groceries | 10.12 (9.50, 10.76) | 64.09 (60.54, 67.50) | 34.93 (32.89, 37.04) | 51.01 (48.04, 53.97) | 34.84 (32.67, 37.09) | 24.02 (22.56, 25.54) |
| Arranging day/night care | 1.46 (1.27, 1.68) | 14.18 (12.46, 16.10) | 5.09 (4.41, 5.87) | 9.00 (7.92, 10.21) | 4.24 (3.57, 5.04) | 3.37 (2.96, 3.84) |
| Getting dressed/undressed | 3.28 (3.00, 3.59) | 40.35 (37.23, 43.56) | 11.72 (10.69, 12.83) | 25.56 (23.44, 27.81) | 9.91 (8.94, 10.97) | 8.20 (7.50, 8.97) |
| Bathing | 3.83 (3.51, 4.19) | 44.99 (41.71, 48.31) | 13.69 (12.51, 14.96) | 29.87 (27.50, 32.36) | 11.52 (10.43, 12.71) | 9.58 (8.74, 10.50) |
| Lying down in bed/rising from bed | 1.46 (1.28, 1.66) | 18.96 (16.73, 21.41) | 5.18 (4.54, 5.91) | 11.31 (9.93, 12.84) | 4.15 (3.55, 4.85) | 3.76 (3.29, 4.29) |
| Eating/drinking, once food has been served or cut | 0.49 (0.40, 0.60) | 6.25 (5.09, 7.65) | 1.72 (1.40, 2.11) | 3.75 (3.06, 4.60) | 1.26 (0.97, 1.63) | 1.26 (1.03, 1.55) |
| Taking medication | 2.78 (2.51, 3.08) | 24.09 (21.66, 26.71) | 9.42 (8.48, 10.45) | 16.15 (14.54, 17.90) | 9.00 (8.01, 10.09) | 6.57 (5.94, 7.26) |
| Using mode of transportation | 5.76 (5.30, 6.26) | 35.80 (32.82, 38.89) | 18.97 (17.49, 20.55) | 28.31 (26.02, 30.71) | 18.16 (16.63, 19.79) | 13.03 (12.02, 14.11) |
| Using computer/tablet/other electronic device | 1.98 (1.64, 2.39) | 9.29 (7.68, 11.20) | 5.71 (4.71, 6.90) | 7.54 (6.00, 9.43) | 5.68 (4.61, 6.98) | 3.62 (3.00, 4.37) |
| Managing medical appointments | 4.55 (4.14, 5.00) | 33.05 (30.12, 36.10) | 15.38 (13.99, 16.89) | 24.45 (22.24, 26.81) | 14.48 (13.03, 16.07) | 10.51 (9.55, 11.55) |
| Preparing meals | 4.25 (3.89, 4.64) | 41.09 (37.89, 44.37) | 14.84 (13.57, 16.21) | 27.62 (25.31, 30.06) | 13.35 (12.08, 14.73) | 10.35 (9.47, 11.30) |
| Moving around all areas of floor | 0.72 (0.60, 0.85) | 8.44 (7.08, 10.04) | 2.52 (2.12, 2.99) | 5.50 (4.64, 6.51) | 1.86 (1.53, 2.26) | 1.83 (1.54, 2.17) |
| Sitting down/standing up from chair | 1.07 (0.92, 1.24) | 13.52 (11.64, 15.65) | 3.80 (3.26, 4.42) | 8.28 (7.13, 9.61) | 2.98 (2.46, 3.59) | 2.72 (2.34, 3.17) |
| Leaving home | 3.45 (3.14, 3.78) | 31.61 (28.76, 34.60) | 12.07 (10.97, 13.25) | 21.58 (19.64, 23.65) | 11.39 (10.26, 12.62) | 8.65 (7.86, 9.52) |
| Executing usual administrative procedures | 7.67 (7.10, 8.29) | 43.12 (39.90, 46.40) | 23.83 (22.08, 25.67) | 34.29 (31.69, 36.98) | 23.71 (21.84, 25.68) | 16.77 (15.49, 18.13) |
| Running everyday errands/tasks in the home | 9.59 (8.98, 10.24) | 62.58 (58.97, 66.05) | 33.65 (31.62, 35.76) | 48.62 (45.70, 51.54) | 33.61 (31.44, 35.85) | 22.74 (21.29, 24.25) |
| Running less frequent tasks | 10.33 (9.66, 11.04) | 50.47 (47.06, 53.87) | 35.34 (33.20, 37.54) | 41.54 (38.82, 44.31) | 35.98 (33.68, 38.35) | 22.68 (21.25, 24.17) |
| Using telephone | 1.28 (1.09, 1.51) | 9.58 (8.16, 11.22) | 4.55 (3.86, 5.36) | 6.78 (5.75, 7.97) | 4.44 (3.71, 5.30) | 3.02 (2.55, 3.56) |
| Using the toilet | 0.81 (0.70, 0.94) | 10.61 (9.14, 12.29) | 2.89 (2.48, 3.36) | 6.31 (5.42, 7.33) | 2.21 (1.84, 2.65) | 2.05 (1.76, 2.38) |
| Choosing appropriate clothes | 1.24 (1.05, 1.45) | 12.54 (10.91, 14.38) | 4.37 (3.72, 5.14) | 8.05 (6.88, 9.40) | 3.69 (3.02, 4.49) | 3.00 (2.57, 3.50) |

**Abbreviations:** GIR: Groupe Isso-Resources, a French measure of autonomy; ADL: activities of daily living; IADL: instrumental activities of daily living

**Table S16. Unmet Need for General Population and Socio-demographic Subgroups**

| | General | Socio-demographic Subgroups | | | |
|---|---|---|---|---|---|
| | | Female | Low-education | Oldest-old | Renounced to care |
| **AP** | 0.52 (0.33, 0.84) | 0.51 (0.25, 1.03) | 0.39 (0.24, 0.63) | 0.35 (0.15, 0.82) | 0.38 (0.17, 0.87) |
| **AP for seeing, reading** | 0.49 (0.29, 0.81) | 0.40 (0.18, 0.85) | 0.43 (0.24, 0.76) | 0.49 (0.17, 1.36) | 0.85 (0.25, 2.79) |
| **AP for hearing, speaking** | 2.94 (2.49, 3.47) | 2.61 (2.02, 3.36) | 3.07 (2.54, 3.72) | 4.45 (3.46, 5.70) | 5.21 (3.65, 7.39) |
| **Prosthetics and implants** | 0.69 (0.49, 0.98) | 0.35 (0.20, 0.59) | 0.82 (0.55, 1.21) | 0.72 (0.35, 1.49) | 1.60 (1.06, 2.42) |
| **Orthoses** | 0.59 (0.41, 0.87) | 0.66 (0.41, 1.06) | 0.55 (0.35, 0.87) | 0.75 (0.43, 1.31) | 1.25 (0.79, 1.97) |
| **AP for personal care** | 0.10 (0.06, 0.16) | 0.11 (0.05, 0.22) | 0.13 (0.07, 0.23) | 0.28 (0.13, 0.59) | 0.48 (0.20, 1.14) |
| **AP for everyday life** | 1.06 (0.80, 1.39) | 1.21 (0.83, 1.77) | 1.10 (0.80, 1.51) | 1.23 (0.76, 1.98) | 4.29 (2.80, 6.52) |
| **AP for personal mobility** | 0.52 (0.35, 0.76) | 0.67 (0.41, 1.09) | 0.73 (0.48, 1.11) | 1.74 (1.09, 2.77) | 0.35 (0.15, 0.82) |
| **AP for transfers, getting up, and going to bed** | 0.21 (0.16, 0.28) | 0.23 (0.16, 0.33) | 0.21 (0.15, 0.30) | 0.55 (0.36, 0.83) | 0.36 (0.18, 0.74) |
| **AP for communication and to manage everyday life** | 0.28 (0.17, 0.44) | 0.43 (0.25, 0.73) | 0.28 (0.16, 0.46) | 0.50 (0.31, 0.79) | 0.57 (0.30, 1.06) |
| **Home Modifications** | 3.57 (3.07, 4.14) | 4.18 (3.44, 5.07) | 4.28 (3.61, 5.07) | 3.60 (2.77, 4.65) | 7.25 (5.54, 9.43) |
| **Toilet Area** | 1.91 (1.59, 2.28) | 2.22 (1.77, 2.78) | 2.26 (1.87, 2.74) | 3.93 (2.94, 5.23) | 3.71 (2.68, 5.10) |
| **Bathing Area** | 4.27 (3.74, 4.88) | 5.03 (4.23, 5.97) | 5.13 (4.43, 5.94) | 5.47 (4.47, 6.69) | 8.45 (6.67, 10.65) |
| **Kitchen** | 0.64 (0.49, 0.83) | 0.69 (0.55, 0.87) | 0.66 (0.48, 0.91) | 0.92 (0.63, 1.32) | 1.66 (1.13, 2.44) |
| **Stairs** | 1.27 (1.04, 1.56) | 1.31 (1.02, 1.68) | 1.47 (1.16, 1.86) | 2.38 (1.76, 3.22) | 2.99 (2.04, 4.35) |
| **Other areas of the residence** | 1.06 (0.86, 1.31) | 1.36 (1.07, 1.73) | 1.24 (0.98, 1.57) | 2.24 (1.68, 2.98) | 2.02 (1.45, 2.81) |
| **Human Help** | 3.08 (2.61, 3.63) | 3.80 (3.09, 4.68) | 3.25 (2.71, 3.90) | 4.66 (3.44, 6.29) | 7.74 (5.70, 10.42) |
| Having regular, balanced diet | 0.25 (0.15, 0.39) | 0.18 (0.12, 0.27) | 0.31 (0.18, 0.54) | 0.38 (0.23, 0.64) | 0.88 (0.55, 1.43) |
| Managing money, budget | 0.14 (0.09, 0.21) | 0.12 (0.07, 0.21) | 0.17 (0.11, 0.28) | 0.25 (0.12, 0.52) | 0.41 (0.19, 0.91) |

| | | | | | |
|---|---|---|---|---|---|
| Finding one's path | 0.12 (0.05, 0.28) | 0.10 (0.06, 0.19) | 0.16 (0.06, 0.42) | 0.26 (0.14, 0.49) | 0.26 (0.10, 0.67) |
| Cutting food/serving drinks | 0.09 (0.05, 0.14) | 0.12 (0.07, 0.21) | 0.11 (0.06, 0.19) | 0.22 (0.11, 0.44) | 0.38 (0.17, 0.86) |
| Doing groceries | 0.90 (0.70, 1.15) | 1.05 (0.79, 1.40) | 1.21 (0.92, 1.60) | 1.57 (1.11, 2.21) | 2.62 (1.48, 4.60) |
| Arranging day/night care | 0.16 (0.11, 0.23) | 0.22 (0.14, 0.35) | 0.22 (0.14, 0.34) | 0.53 (0.31, 0.89) | 0.61 (0.26, 1.39) |
| Getting dressed/undressed | 0.16 (0.10, 0.27) | 0.25 (0.14, 0.44) | 0.20 (0.11, 0.37) | 0.44 (0.28, 0.68) | 0.22 (0.09, 0.56) |
| Bathing | 0.29 (0.18, 0.45) | 0.33 (0.21, 0.53) | 0.37 (0.22, 0.62) | 0.89 (0.48, 1.63) | 0.88 (0.28, 2.75) |
| Lying down in bed/rising from bed | 0.06 (0.04, 0.11) | 0.08 (0.04, 0.16) | 0.08 (0.04, 0.14) | 0.13 (0.06, 0.30) | 0.24 (0.09, 0.61) |
| Eating/drinking, once food has been served or cut | 0.04 (0.02, 0.08) | 0.04 (0.02, 0.11) | 0.05 (0.02, 0.12) | 0.11 (0.04, 0.31) | 0.09 (0.02, 0.38) |
| Taking medication | 0.13 (0.06, 0.29) | 0.07 (0.03, 0.17) | 0.17 (0.07, 0.43) | 0.18 (0.08, 0.41) | 0.12 (0.03, 0.50) |
| Using mode of transportation | 0.47 (0.34, 0.66) | 0.60 (0.41, 0.88) | 0.54 (0.39, 0.75) | 0.76 (0.51, 1.13) | 1.72 (0.95, 3.10) |
| Using computer/tablet/other electronic device | 1.35 (1.02, 1.77) | 1.72 (1.21, 2.43) | 1.50 (1.11, 2.01) | 2.17 (1.32, 3.57) | 2.15 (1.26, 3.65) |
| Managing medical appointments | 0.24 (0.17, 0.35) | 0.27 (0.17, 0.43) | 0.30 (0.21, 0.43) | 0.56 (0.31, 0.99) | 0.76 (0.43, 1.36) |
| Preparing meals | 0.40 (0.27, 0.59) | 0.48 (0.28, 0.82) | 0.46 (0.31, 0.67) | 1.05 (0.61, 1.80) | 0.94 (0.57, 1.53) |
| Moving around all areas of floor | 0.07 (0.02, 0.20) | 0.12 (0.04, 0.36) | 0.10 (0.03, 0.31) | 0.12 (0.04, 0.31) | 0.16 (0.05, 0.46) |
| Sitting down/standing up from chair | 0.07 (0.02, 0.18) | 0.11 (0.03, 0.33) | 0.10 (0.03, 0.28) | 0.28 (0.08, 0.95) | 0.21 (0.09, 0.52) |
| Leaving home | 0.21 (0.15, 0.29) | 0.26 (0.18, 0.37) | 0.27 (0.19, 0.37) | 0.60 (0.39, 0.93) | 0.54 (0.28, 1.05) |
| Executing usual administrative procedures | 1.21 (0.92, 1.59) | 1.16 (0.82, 1.62) | 1.42 (1.06, 1.91) | 1.49 (0.92, 2.42) | 2.99 (1.82, 4.85) |
| Running everyday errands/tasks in the home | 2.33 (1.99, 2.73) | 2.90 (2.39, 3.52) | 2.53 (2.12, 3.01) | 4.67 (3.70, 5.88) | 5.73 (4.13, 7.91) |
| Running less frequent tasks | 2.01 (1.63, 2.47) | 2.65 (2.08, 3.37) | 2.02 (1.60, 2.53) | 3.41 (2.36, 4.90) | 3.85 (2.69, 5.49) |
| Using telephone | 0.20 (0.10, 0.41) | 0.20 (0.08, 0.52) | 0.20 (0.09, 0.43) | 0.32 (0.18, 0.57) | 0.28 (0.13, 0.62) |
| Using the toilet | 0.03 (0.02, 0.07) | 0.04 (0.02, 0.11) | 0.05 (0.02, 0.10) | 0.12 (0.04, 0.31) | 0.16 (0.06, 0.42) |
| Choosing appropriate clothes | 0.12 (0.05, 0.24) | 0.09 (0.03, 0.26) | 0.16 (0.07, 0.36) | 0.38 (0.13, 1.14) | 0.13 (0.04, 0.44) |

**Table S17. Unmet Need for General Population and Morbidity Subgroups**

| | General | Morbidity Subgroups | | |
|---|---|---|---|---|
| | | Frail | Any Chronic Condition | Multimorbid |
| **AP** | 0.52 (0.33, 0.84) | 0.84 (0.47, 1.49) | 0.49 (0.30, 0.82) | 0.46 (0.26, 0.82) |
| **AP for seeing, reading** | 0.49 (0.29, 0.81) | 0.77 (0.42, 1.43) | 0.48 (0.28, 0.83) | 0.29 (0.14, 0.58) |
| **AP for hearing, speaking** | 2.94 (2.49, 3.47) | 4.21 (3.45, 5.11) | 3.26 (2.74, 3.87) | 3.61 (2.99, 4.36) |
| **Prosthetics and implants** | 0.69 (0.49, 0.98) | 1.11 (0.74, 1.66) | 0.83 (0.58, 1.18) | 0.84 (0.57, 1.24) |
| **Orthoses** | 0.59 (0.41, 0.87) | 1.19 (0.87, 1.62) | 0.74 (0.51, 1.09) | 0.90 (0.60, 1.35) |
| **AP for personal care** | 0.10 (0.06, 0.16) | 0.36 (0.21, 0.62) | 0.12 (0.07, 0.20) | 0.13 (0.07, 0.24) |
| **AP for everyday life** | 1.06 (0.80, 1.39) | 2.21 (1.65, 2.96) | 1.26 (0.95, 1.66) | 1.68 (1.26, 2.25) |
| **AP for personal mobility** | 0.52 (0.35, 0.76) | 1.55 (1.07, 2.26) | 0.61 (0.41, 0.91) | 0.77 (0.51, 1.15) |
| **AP for transfers, getting up, and going to bed** | 0.21 (0.16, 0.28) | 0.80 (0.60, 1.08) | 0.26 (0.20, 0.35) | 0.34 (0.25, 0.46) |
| **AP for communication and to manage everyday life** | 0.28 (0.17, 0.44) | 0.70 (0.50, 0.98) | 0.35 (0.22, 0.55) | 0.46 (0.28, 0.74) |
| **Home Modifications** | 3.57 (3.07, 4.14) | 8.53 (7.31, 9.94) | 4.44 (3.82, 5.15) | 5.79 (4.96, 6.75) |
| **Toilet Area** | 1.91 (1.59, 2.28) | 5.40 (4.54, 6.41) | 2.31 (1.93, 2.76) | 3.05 (2.53, 3.67) |
| **Bathing Area** | 4.27 (3.74, 4.88) | 10.55 (9.26, 12.00) | 5.30 (4.64, 6.05) | 7.01 (6.12, 8.04) |
| **Kitchen** | 0.64 (0.49, 0.83) | 1.94 (1.51, 2.48) | 0.80 (0.61, 1.04) | 0.98 (0.75, 1.27) |
| **Stairs** | 1.27 (1.04, 1.56) | 3.59 (2.90, 4.43) | 1.60 (1.30, 1.96) | 2.14 (1.74, 2.64) |
| **Other areas of the residence** | 1.06 (0.86, 1.31) | 3.29 (2.70, 4.00) | 1.33 (1.08, 1.64) | 1.76 (1.42, 2.18) |
| **Human Help** | 3.08 (2.61, 3.63) | 7.53 (6.29, 9.01) | 3.77 (3.19, 4.44) | 4.56 (3.82, 5.44) |
| Having regular, balanced diet | 0.25 (0.15, 0.39) | 0.93 (0.57, 1.51) | 0.31 (0.19, 0.49) | 0.39 (0.24, 0.65) |
| Managing money, budget | 0.14 (0.09, 0.21) | 0.34 (0.20, 0.56) | 0.17 (0.11, 0.25) | 0.20 (0.13, 0.32) |

| | | | | |
|---|---|---|---|---|
| Finding one's path | 0.12 (0.05, 0.28) | 0.48 (0.20, 1.12) | 0.15 (0.06, 0.35) | 0.20 (0.08, 0.49) |
| Cutting food/serving drinks | 0.09 (0.05, 0.14) | 0.35 (0.22, 0.56) | 0.11 (0.06, 0.17) | 0.13 (0.08, 0.22) |
| Doing groceries | 0.90 (0.70, 1.15) | 2.65 (2.05, 3.42) | 1.12 (0.87, 1.44) | 1.49 (1.14, 1.93) |
| Arranging day/night care | 0.16 (0.11, 0.23) | 0.61 (0.41, 0.92) | 0.20 (0.13, 0.29) | 0.28 (0.19, 0.42) |
| Getting dressed/undressed | 0.16 (0.10, 0.27) | 0.60 (0.35, 1.03) | 0.20 (0.12, 0.33) | 0.20 (0.13, 0.29) |
| Bathing | 0.29 (0.18, 0.45) | 0.89 (0.59, 1.33) | 0.36 (0.23, 0.56) | 0.34 (0.25, 0.47) |
| Lying down in bed/rising from bed | 0.06 (0.04, 0.11) | 0.23 (0.14, 0.40) | 0.08 (0.05, 0.14) | 0.11 (0.07, 0.19) |
| Eating/drinking, once food has been served or cut | 0.04 (0.02, 0.08) | 0.15 (0.07, 0.32) | 0.05 (0.02, 0.10) | 0.06 (0.03, 0.14) |
| Taking medication | 0.13 (0.06, 0.29) | 0.50 (0.22, 1.15) | 0.16 (0.07, 0.36) | 0.22 (0.09, 0.51) |
| Using mode of transportation | 0.47 (0.34, 0.66) | 1.33 (1.03, 1.71) | 0.59 (0.42, 0.82) | 0.68 (0.50, 0.92) |
| Using computer/tablet/other electronic device | 1.35 (1.02, 1.77) | 2.62 (1.89, 3.61) | 1.64 (1.24, 2.17) | 2.06 (1.54, 2.77) |
| Managing medical appointments | 0.24 (0.17, 0.35) | 0.77 (0.51, 1.15) | 0.30 (0.21, 0.43) | 0.36 (0.24, 0.53) |
| Preparing meals | 0.40 (0.27, 0.59) | 1.20 (0.85, 1.70) | 0.50 (0.34, 0.73) | 0.67 (0.45, 1.01) |
| Moving around all areas of floor | 0.07 (0.02, 0.20) | 0.27 (0.09, 0.80) | 0.08 (0.03, 0.25) | 0.11 (0.04, 0.35) |
| Sitting down/standing up from chair | 0.07 (0.02, 0.18) | 0.25 (0.09, 0.73) | 0.08 (0.03, 0.23) | 0.11 (0.04, 0.32) |
| Leaving home | 0.21 (0.15, 0.29) | 0.79 (0.57, 1.08) | 0.26 (0.19, 0.36) | 0.31 (0.22, 0.43) |
| Executing usual administrative procedures | 1.21 (0.92, 1.59) | 2.49 (1.87, 3.31) | 1.50 (1.14, 1.97) | 1.91 (1.42, 2.55) |
| Running everyday errands/tasks in the home | 2.33 (1.99, 2.73) | 6.87 (5.79, 8.14) | 2.86 (2.44, 3.36) | 3.70 (3.12, 4.37) |
| Running less frequent tasks | 2.01 (1.63, 2.47) | 5.36 (4.33, 6.62) | 2.50 (2.03, 3.08) | 3.19 (2.57, 3.96) |
| Using telephone | 0.20 (0.10, 0.41) | 0.50 (0.22, 1.13) | 0.25 (0.12, 0.52) | 0.26 (0.13, 0.53) |
| Using the toilet | 0.03 (0.02, 0.07) | 0.13 (0.06, 0.28) | 0.04 (0.02, 0.09) | 0.06 (0.03, 0.12) |
| Choosing appropriate clothes | 0.12 (0.05, 0.24) | 0.32 (0.15, 0.66) | 0.14 (0.07, 0.30) | 0.15 (0.07, 0.30) |

**Table S18. Unmet Need for General Population and Limitations Subgroups**

| | General | Limitations Subgroups | | | | |
|---|---|---|---|---|---|---|
| | | GIR | ADL/IADL | ADL | IADL | Self-reported limitations |
| **AP** | 0.52 (0.33, 0.84) | 0.83 (0.45, 1.54) | 0.57 (0.38, 0.84) | 0.74 (0.45, 1.22) | 0.59 (0.40, 0.89) | 0.83 (0.48, 1.42) |
| **AP for seeing, reading** | 0.49 (0.29, 0.81) | 0.70 (0.38, 1.30) | 0.61 (0.33, 1.14) | 0.87 (0.36, 2.09) | 0.68 (0.37, 1.26) | 0.76 (0.40, 1.43) |
| **AP for hearing, speaking** | 2.94 (2.49, 3.47) | 5.65 (4.34, 7.32) | 4.32 (3.49, 5.33) | 4.80 (3.78, 6.08) | 4.26 (3.40, 5.32) | 3.75 (3.09, 4.53) |
| **Prosthetics and implants** | 0.69 (0.49, 0.98) | 1.16 (0.67, 1.97) | 1.01 (0.68, 1.49) | 1.58 (0.97, 2.57) | 0.89 (0.63, 1.24) | 1.15 (0.77, 1.73) |
| **Orthoses** | 0.59 (0.41, 0.87) | 1.48 (0.82, 2.67) | 1.20 (0.81, 1.77) | 1.51 (1.01, 2.25) | 1.20 (0.79, 1.83) | 1.00 (0.68, 1.47) |
| **AP for personal care** | 0.10 (0.06, 0.16) | 0.71 (0.34, 1.48) | 0.30 (0.17, 0.52) | 0.52 (0.27, 0.98) | 0.31 (0.17, 0.55) | 0.25 (0.15, 0.42) |
| **AP for everyday life** | 1.06 (0.80, 1.39) | 4.02 (2.58, 6.22) | 2.35 (1.76, 3.15) | 3.95 (2.78, 5.57) | 2.26 (1.65, 3.07) | 2.01 (1.50, 2.70) |
| **AP for personal mobility** | 0.52 (0.35, 0.76) | 1.40 (0.91, 2.15) | 1.65 (1.10, 2.46) | 1.83 (1.18, 2.82) | 1.73 (1.14, 2.63) | 1.29 (0.86, 1.93) |
| **AP for transfers, getting up, and going to bed** | 0.21 (0.16, 0.28) | 1.62 (1.13, 2.32) | 0.70 (0.52, 0.94) | 1.22 (0.88, 1.70) | 0.68 (0.49, 0.93) | 0.53 (0.40, 0.72) |
| **AP for communication and to manage everyday life** | 0.28 (0.17, 0.44) | 1.15 (0.75, 1.77) | 0.68 (0.49, 0.93) | 0.80 (0.53, 1.18) | 0.69 (0.49, 0.97) | 0.58 (0.37, 0.90) |
| **Home Modifications** | 3.57 (3.07, 4.14) | 11.19 (9.00, 13.84) | 8.77 (7.45, 10.29) | 10.11 (8.41, 12.12) | 9.15 (7.72, 10.82) | 7.01 (6.04, 8.13) |
| **Toilet Area** | 1.91 (1.59, 2.28) | 9.96 (8.02, 12.29) | 5.50 (4.58, 6.60) | 8.34 (6.80, 10.19) | 5.83 (4.82, 7.03) | 4.46 (3.71, 5.36) |
| **Bathing Area** | 4.27 (3.74, 4.88) | 14.84 (12.42, 17.64) | 10.62 (9.26, 12.16) | 13.29 (11.47, 15.34) | 10.99 (9.51, 12.66) | 8.52 (7.46, 9.71) |
| **Kitchen** | 0.64 (0.49, 0.83) | 3.86 (2.99, 4.98) | 1.83 (1.43, 2.32) | 3.10 (2.35, 4.09) | 1.94 (1.52, 2.49) | 1.36 (1.07, 1.72) |
| **Stairs** | 1.27 (1.04, 1.56) | 6.03 (4.53, 7.99) | 3.40 (2.74, 4.21) | 5.12 (4.01, 6.53) | 3.46 (2.75, 4.34) | 2.83 (2.30, 3.49) |
| **Other areas of the residence** | 1.06 (0.86, 1.31) | 7.11 (5.36, 9.37) | 3.33 (2.70, 4.11) | 5.45 (4.25, 6.98) | 3.44 (2.75, 4.29) | 2.55 (2.06, 3.14) |
| **Human Help** | 3.08 (2.61, 3.63) | 7.51 (5.43, 10.29) | 9.94 (8.40, 11.73) | 9.10 (7.16, 11.50) | 10.40 (8.75, 12.32) | 5.89 (4.96, 6.99) |
| Having regular, balanced diet | 0.25 (0.15, 0.39) | 2.02 (1.02, 3.93) | 0.88 (0.55, 1.40) | 1.47 (0.83, 2.59) | 0.90 (0.55, 1.48) | 0.53 (0.31, 0.91) |
| Managing money, budget | 0.14 (0.09, 0.21) | 0.27 (0.12, 0.61) | 0.49 (0.32, 0.74) | 0.47 (0.26, 0.84) | 0.48 (0.31, 0.76) | 0.25 (0.16, 0.39) |

| | | | | | | |
|---|---|---|---|---|---|---|
| Finding one's path | 0.12 (0.05, 0.28) | 1.21 (0.42, 3.48) | 0.42 (0.18, 1.00) | 0.76 (0.28, 2.10) | 0.47 (0.20, 1.10) | 0.31 (0.13, 0.73) |
| Cutting food/serving drinks | 0.09 (0.05, 0.14) | 0.86 (0.51, 1.45) | 0.31 (0.19, 0.50) | 0.54 (0.33, 0.90) | 0.33 (0.20, 0.53) | 0.21 (0.13, 0.35) |
| Doing groceries | 0.90 (0.70, 1.15) | 3.51 (2.29, 5.34) | 3.09 (2.39, 3.98) | 3.46 (2.47, 4.82) | 3.29 (2.53, 4.28) | 1.81 (1.40, 2.35) |
| Arranging day/night care | 0.16 (0.11, 0.23) | 1.94 (1.28, 2.93) | 0.54 (0.36, 0.81) | 1.13 (0.74, 1.71) | 0.53 (0.34, 0.82) | 0.41 (0.28, 0.62) |
| Getting dressed/undressed | 0.16 (0.10, 0.27) | 1.94 (1.13, 3.30) | 0.57 (0.35, 0.95) | 1.18 (0.70, 2.00) | 0.56 (0.32, 0.97) | 0.40 (0.23, 0.67) |
| Bathing | 0.29 (0.18, 0.45) | 2.24 (1.37, 3.63) | 1.02 (0.65, 1.60) | 2.03 (1.25, 3.29) | 1.07 (0.66, 1.72) | 0.69 (0.43, 1.12) |
| Lying down in bed/rising from bed | 0.06 (0.04, 0.11) | 0.68 (0.39, 1.17) | 0.23 (0.14, 0.39) | 0.45 (0.26, 0.77) | 0.19 (0.10, 0.35) | 0.15 (0.08, 0.26) |
| Eating/drinking, once food has been served or cut | 0.04 (0.02, 0.08) | 0.47 (0.22, 1.04) | 0.13 (0.06, 0.28) | 0.28 (0.13, 0.62) | 0.14 (0.06, 0.31) | 0.09 (0.04, 0.21) |
| Taking medication | 0.13 (0.06, 0.29) | 1.15 (0.37, 3.47) | 0.46 (0.20, 1.03) | 0.73 (0.25, 2.08) | 0.48 (0.20, 1.13) | 0.34 (0.15, 0.76) |
| Using mode of transportation | 0.47 (0.34, 0.66) | 2.32 (1.65, 3.25) | 1.64 (1.16, 2.30) | 1.82 (1.35, 2.45) | 1.81 (1.29, 2.54) | 0.99 (0.70, 1.39) |
| Using computer/tablet/other electronic device | 1.35 (1.02, 1.77) | 2.69 (1.57, 4.56) | 4.31 (3.23, 5.74) | 3.91 (2.59, 5.85) | 4.41 (3.27, 5.92) | 2.43 (1.81, 3.25) |
| Managing medical appointments | 0.24 (0.17, 0.35) | 0.94 (0.46, 1.91) | 0.76 (0.52, 1.12) | 0.76 (0.43, 1.35) | 0.80 (0.54, 1.20) | 0.52 (0.35, 0.77) |
| Preparing meals | 0.40 (0.27, 0.59) | 2.84 (1.68, 4.76) | 1.42 (0.96, 2.09) | 2.38 (1.53, 3.68) | 1.43 (0.94, 2.16) | 0.92 (0.60, 1.41) |
| Moving around all areas of floor | 0.07 (0.02, 0.20) | 0.42 (0.19, 0.89) | 0.24 (0.08, 0.71) | 0.25 (0.12, 0.53) | 0.27 (0.09, 0.79) | 0.18 (0.06, 0.52) |
| Sitting down/standing up from chair | 0.07 (0.02, 0.18) | 0.41 (0.20, 0.83) | 0.23 (0.08, 0.65) | 0.26 (0.13, 0.50) | 0.26 (0.09, 0.72) | 0.17 (0.06, 0.48) |
| Leaving home | 0.21 (0.15, 0.29) | 1.46 (0.97, 2.18) | 0.75 (0.55, 1.03) | 1.17 (0.83, 1.64) | 0.83 (0.61, 1.13) | 0.55 (0.40, 0.75) |
| Executing usual administrative procedures | 1.21 (0.92, 1.59) | 3.61 (2.12, 6.09) | 3.83 (2.87, 5.10) | 2.95 (1.96, 4.41) | 4.08 (3.02, 5.48) | 2.07 (1.55, 2.76) |
| Running everyday errands/tasks in the home | 2.33 (1.99, 2.73) | 6.47 (4.93, 8.45) | 7.79 (6.65, 9.11) | 7.36 (5.93, 9.09) | 8.40 (7.15, 9.85) | 5.20 (4.40, 6.15) |
| Running less frequent tasks | 2.01 (1.63, 2.47) | 6.67 (4.78, 9.24) | 6.99 (5.68, 8.56) | 6.83 (5.24, 8.86) | 7.59 (6.16, 9.32) | 4.44 (3.59, 5.49) |
| Using telephone | 0.20 (0.10, 0.41) | 1.15 (0.38, 3.45) | 0.68 (0.32, 1.44) | 0.91 (0.38, 2.15) | 0.74 (0.35, 1.59) | 0.51 (0.24, 1.07) |
| Using the toilet | 0.03 (0.02, 0.07) | 0.38 (0.17, 0.86) | 0.12 (0.06, 0.25) | 0.26 (0.12, 0.55) | 0.13 (0.06, 0.28) | 0.09 (0.04, 0.19) |
| Choosing appropriate clothes | 0.12 (0.05, 0.24) | 0.23 (0.09, 0.61) | 0.27 (0.13, 0.57) | 0.25 (0.12, 0.50) | 0.30 (0.14, 0.63) | 0.21 (0.10, 0.43) |

**Abbreviations:** GIR: Groupe Isso-Resources, a French measure of autonomy; ADL: activities of daily living; IADL: instrumental activities of daily living

**Figure S1. List of Assistive Technologies**

Participants reported their use and need for 54 assistive products across nine functional groups, 34 home modifications across five living areas and 24 activities requiring human help

### Assistive Products (AP):

1. AP for seeing, reading
   - Glasses/contact lenses
   - Optical/electronic magnifying glass
   - Electronic magnifier
   - Vocal recognition system
   - Character reading machines, reading machines
   - Other product for seeing, reading
2. AP for hearing, speaking
   - Hearing implants
   - Hearing aids
   - Material to replace the sounds of the home
   - AV material for the deaf, hard of hearing
   - Induction/hearing loop
   - Voice generator/amplifier, communication amplifiers
   - Other product for hearing/speaking
3. Prosthetics and implants
   - Upper limb prosthetics/orthoses
   - Lower limb prostheses/othoses
   - Other prostheses
4. Orthoses
   - Lower limb orthoses
   - Upper limb orthoses
   - Spinal orthoses
   - Other orthoses
5. AP for communication, to manage everyday life
   - Accessible input devices for computers/tablets
   - Adapted fixed network telephone
   - Adapted mobile network telephone
   - Connected object
   - Connected speakers
   - Input software/application
   - Classic/adapted remote controls to open home/manage lights
   - Device also used for other things
6. AP for everyday life
   - Dentures
   - AP for washing, bathing, showering
   - AP for dressing
   - AP for eating, drinking
   - AP for toileting
   - AP for domestic activities
   - AP for getting in and out of bed
   - Personal emergency alarm systen
   - Other products for everyday life
7. AP for personal mobility
   - Canes, walking sticks
   - White cane
   - Walker
   - Manual wheelchair
   - Electric wheelchair
   - Tricycle, adapted bicycle, adapted scooter
   - Help from an animal
   - Other products for personal mobility
8. AP for changing position
   - Patient lift, not fixed to the ceiling/walls
   - Transfer board, lifting strap
   - Device to facilitate standing transfer
   - Other products for changing position
9. AP for personal care
   - Catheter/device for collecting urine
   - Absorbing products
   - Adapted clothes, shoes
   - AP for ostomy care
   - AP to manage tissue integrity

### Home Modifications:

1. Toilet Area
   - Supporting handrails, grab bars
   - Raised toilet seats/seats with built-in raising mechanism
   - Floors with adapted materials
   - Accessible electrical outlet/switch
   - Area with enough space
   - Single area for toilet, bathing areas
2. Bathing Area
   - Supporting handrails, grab bars
   - Enlarged shower unit/open shower stall
   - Bath/shower seat fixed to the wall
   - Accessible sink
   - Floors with adapted materials
   - Accessible electrical outlet/switch
   - Area with enough space
   - Single area for toilet, bathing areas
3. Stairs
   - One/two supporting handrails
   - Stairlifts with seat
   - Platform lift
   - Elevator
4. Kitchen
   - Counters, lowered/height-adjustable
   - Accessible sink, lowered/height-adjustable
   - Specialized furniture
   - Specialized equipment
   - Ramp/handrail/grab bar
   - Floors with adapted materials
   - Accessible electrical outlet/switch
5. Other areas of the residence
   - Stationary hoists fixed to ceiling/walls
   - Supporting handrails, grab bars
   - Automatic light fixtures/light strip
   - Light device to alert to noises
   - Motorized windows/electric curtains
   - Automatic doors
   - Enlarged doors/hallways
   - Non-slip floor coverings
   - Other furniture, lowered/height-adjustable

### Human Help:

1. Bathing
2. Getting dressed/undressed
3. Cutting food or helping yourself to drinks
4. Eating/drinking, once food has been served or cut
5. Using the toilet
6. Lying down in bed/rising from bed
7. Sitting down/standing up from chair
8. Shopping for groceries
9. Preparing meals
10. For common household chores
11. For undertaking less frequent tasks
12. To carry out routine administrative procedures
13. Taking medication
14. Moving around all areas of floor
15. Leaving home
16. Using a mode of transportation
17. Finding your way when you go out
18. Using a telephone
19. Using computer/tablet/other electronic device
20. Choosing clothes adapted to circumstances
21. Having a regular, balanced diet
22. Managing money, budget
23. Managing medical appointments
24. Arranging day/night care

**Figure S2. Proportion of Unmet, Undermet and Met Need among those with Need for the particular category across Assistive Products, Home Modifications and Human Help**

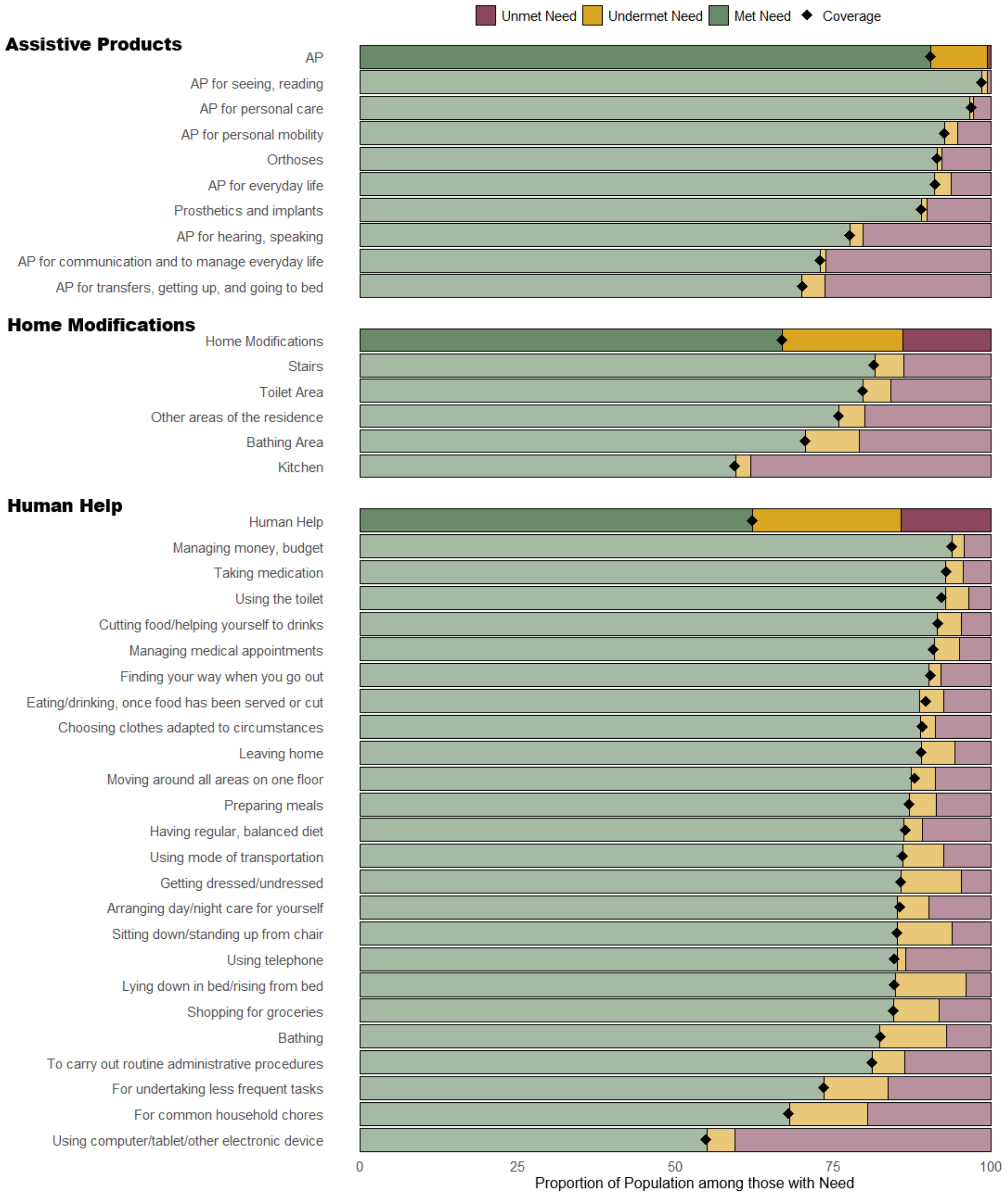


**Abbreviations**: AP: Assistive Products

The population of interest here is those with need so met need is equivalent to coverage.

Met Need: the proportion of the population whose need is fully satisfied by the tool they use, Undermet Need: the proportion of the population that use a tool but report needing an updated or different tool within the same group category, Unmet Need: the proportion of the population who reports needing a tool but does not use one
